# Computational Transformation of Chemical Biology for Precision Therapeutics: Facilitating In-Silico Study of Role of Cuproptosis in Early Detection of Alzheimer’s Disease

**DOI:** 10.64898/2026.05.18.26353543

**Authors:** Pratibha Singh, Soumya Lipsa Rath

## Abstract

**Background:** Alzheimer’s disease (AD) is a multifactorial neurodegenerative disorder in which copper dyshomeostasis, mitochondrial stress, oxidative injury and immune dysregulation may contribute to pathogenesis. Cuproptosis, a copper-triggered regulated cell death pathway, has emerged as a potential mechanistic link to AD, but its therapeutic and biomarker implications remain incompletely defined.

**Methods:** We integrated transcriptomic, machine learning, immune infiltration, QSFR, molecular docking, docking validation and ADME analyses using GEO blood- and brain-based AD cohorts. Differentially expressed genes were intersected with curated cuproptosis-related genes, followed by pathway enrichment, construction and validation of a hybrid ensemble classifier, CIBERSORT-based immune correlation analysis, QSFR-driven target prioritization, ligand docking, consensus docking validation and SwissADME profiling.

**Results:** The transcriptomic analyses revealed reproducible AD associated signatures enriched in neurodegenerative, oxidative stress, mitochondrial and inflammatory pathways. Across multiple machine learning models, FDX1, PDHB, PDHA1, DLAT and DLD consistently emerged as the most important cuproptosis-related genes, with the hybrid ensemble achieving the best diagnostic performance. Immune profiling suggested that these genes are linked to distinct immune infiltration patterns. QSFR and docking prioritized FDX1 as a key target and Clioquinol, PBT2 and Ebselen showed the strongest and most consistent binding behavior. Docking validation confirmed reliable pose reproduction and enrichment over decoys, while ADME analysis supported Clioquinol, PBT2 and Ebselen as the most balanced candidates for further consideration.

**Conclusion:** This integrated workflow identifies a cuproptosis-centered mitochondrial gene module as a robust AD signature and highlights Clioquinol, PBT2 and Ebselen as promising repurposing candidates. The findings provide a prioritized computational framework for future experimental validation of copper-linked therapeutic strategies in AD.

## INTRODUCTION

Alzheimer’s disease (AD) is a progressive neurodegenerative disorder that represents a major global health challenge, affecting millions worldwide with prevalence expected to rise in the coming decades [1]. It is a multifactorial complex neurodegeneartive disorder and is characterized by a progressive cognitive decline, memory impairment and behavioral changes and an accelerated neurodegenerative course, driven by distinct genetic etiologies (such as PSEN1 or APP mutations) and heightened therapeutic vulnerability [2]. The pathological features of AD include synaptic dysfunction, dysregulated autophagy, chronic neuroinflammation, excessive production of reactive oxygen species (ROS), neuronal loss and oxidative stress resulting from disrupted metal homeostasis, alongside the core pathologies including amyloid-β (Aβ) fibrillogenesis and hyperphosphorylated tau tangles. All of these are increasingly recognized to intersect with disturbances in transition metal homeostasis [2,3]. One such transition metal is copper which regulates some of the critical brain functions like energy metabolism, antioxidant defense, neurotransmission, synaptic plasticity and neuronal development. Recent studies have shown that the copper (Cu) dyshomeostasis has emerged as a key feature of AD brain pathology, exhibiting 2- to 5-fold elevations of Cu in amyloid plaques and neuronal compartments [4]. These Cu²⁺ accumulations catalyze Aβ aggregation through Cu²⁺-Aβ coordination at His6, His13 or His14 residues and promote oxidative damage via Fenton-like reactions, thereby amplifying neuronal vulnerability [5].

Emerging paradigms have highlighted cuproptosis, a copper-triggered, non-apoptotic regulated cell death mechanism, as a central process at the chemical-biological interface in EOAD pathogenesis [6]. Distinct from canonical ferroptosis (driven by Fe²⁺-mediated lipid peroxidation), apoptosis or necroptosis, cuproptosis arises from Cu²⁺ overload disrupting mitochondrial bioenergetics at the tricarboxylic acid (TCA) cycle interface [5]. Excess Cu²⁺ (typically more than10 µM intracellularly) preferentially binds dithiolane rings of lipoylated proteins in the pyruvate dehydrogenase (PDH) and α-ketoglutarate dehydrogenase (KGDH) complexes, inducing thiolate aggregation and proteotoxic stress [7]. This triggers ferredoxin 1 (FDX1) dependent mitochondrial Fe-S cluster collapse, halting lipoic acid (LA) biosynthesis, oligomerization of Pyruvate dehydrogenase complex (PDC) and finally TCA flux attenuation leading to a significant reduction in NADH and FADH₂ output [8]. In EOAD neurons, CRGs (cuproptosis related genes) are assumed to exhibit upregulation, spatially colocalizing with Aβ plaques and ultimately correlating with synaptic failure and cognitive decline [6,8].

Current therapeutic strategies for EOAD remain limited as anti-Aβ monoclonal antibodies produce only modest reductions in CDR-SB (Clinical Dementia Rating-Sum of Boxes) progression, whereas metal chelators have failed in clinical trials due to poor specificity and off-target toxicity [9,10]. Advancing precision therapeutics requires a reconfiguration of chemical biology frameworks, integrating computational approaches to systematically interrogate Cu-protein binding landscapes, protein-protein interaction (PPI) dynamics and modulator pharmacophores. Several key gaps constrain progress in the field. EOAD subtype heterogeneity is frequently neglected, as temporally distinct trajectories such as those in PSEN1 associated versus sporadic EOAD remains hidden in bulk analyses [2]. Current models often do not properly link CRGs and Cu, treating CRG expression as independent of Cu-binding free energies, while immune profiling is limited to static snapshots that fail to capture variation across immune cell subpopulations [11,12]. Machine learning approaches are also fragmented, with some methods prioritizing tabular time-series data and others focusing exclusively on graph structured information [13]. Furthermore, translational efforts are hampered by the absence of ADMET-filtered drug repurposing pipelines [14]. Prior work linking cuproptosis to AD has largely relied on bulk RNA-seq or static STRING networks, with limited integration of temporally stratified cross validation [15].

This study aims to catalyze a computational transformation of chemical biology at the interface of redox and systems cell biology by establishing a mechanistically grounded, integrative multi-scale in silico framework for AD linked cuproptosis therapeutics. By integrating multi-omics study including CRG-immune deconvolution, QSFR based Cu²⁺-affinity modeling and a hybrid machine-learning ensemble, it offers a data-driven pipeline to identify and prioritize repurposable candidates that modulate copper-dependent cell-death pathways in neurons. The framework moves beyond static network analyses toward temporally stratified, chemistry-informed models that can guide the development of precision interventions targeting AD linked cuproptosis, while narrowing translational gaps through ADMET-filtered drug repurposing strategies.

## METHODOLOGY

### 1. DATA SET COLLECTION

The Gene Expression Omnibus (GEO) database was used to download the AD datasets [16]. A total of 35 cuproptosis-related genes (CRGs) were curated for this study. This list was compiled by integrating the canonical core regulators with additional regulatory, transport and copper-binding genes identified through a systematic literature search and the databases like GeneCards [17], Kyoto Encyclopedia of Genes and Genomes (KEGG) [18] and Gene Set Enrichment Analysis (GSEA) [19] to ensure a comprehensive assessment of the cuproptosis pathway.

#### 1.1. Dataset Partitioning and Validation

To strengthen the reliability of the analysis, *GSE63060* and *GSE63061* were selected as discovery and training datasets whereas *GSE48350* was used for internal validation. *GSE109887* and *GSE122063* were employed as external validation cohorts and *GSE67333* was used for biological confirmation in brain RNA-seq data. This dataset partitioning enabled stepwise assessment of the robustness, reproducibility and biological relevance of the identified signatures across blood and brain-based cohorts. Table 1 gives summary of the datasets used according to their source tissue, analytical purpose and stage of confirmation.

**Table 1:** Summary of datasets used.

| DATASET | PLATFORM | DESCRIPTION | SOURCE &<br>SAMPLES<br>(Disease/Control) | PURPOSE | REFERENCE |
| --- | --- | --- | --- | --- | --- |
| *GSE63060* | GPL6801 | Blood-based microarray data from the AddNeuroMed cohort, including AD, mild cognitive impairment (MCI) and control samples | Blood<br>145/104 | Discovery & Training | [20] |
| *GSE63061* | GPL6887 | Blood-based microarray data from the AddNeuroMed cohort, including AD, mild cognitive impairment (MCI) and control samples | Blood<br>139/135 | Discovery & Training | [20] |
| *GSE48350* | GPL570 | Brain-region microarray data from normal controls and AD cases across hippocampus, entorhinal cortex, superior frontal cortex and post-central gyrus. | Brain tissue<br>80/173 | Internal Validation | [21-26] |
| *GSE109887* | GPL10904 | Middle temporal gyrus brain expression dataset linked to an AD DNA methylation/hydroxy methylation study, used to connect epigenetic changes with gene expression. | Brain tissue<br>46/32 | External<br>Validation | [27,28] |
| *GSE122063* | GPL570 | Frontal and temporal cortex expression profiling from AD, vascular dementia, and control brain tissues. | Brain tissue<br>12/11 | External<br>Validation | [29] |
| *GSE67333* | RNA-seq | RNA-sequencing dataset from human hippocampus samples that provides a transcriptomic profile of onset of AD compared with age-matched controls. | RNA-seq brain<br>dataset<br>4/4 | Biological<br>Confirmatio<br>n | [30] |

### 2. DIFFERENTIAL EXPRESSION ANALYSIS USING Z-SCORE NORMALIZATION AND FDR CORRECTION

Raw gene expression data from the datasets were retrieved from the NCBI GEO using the GEOquery package in R [31]. Expression matrices were extracted and preprocessed to ensure consistent gene annotation and sample alignment. To standardize expression profiles across samples, z-score normalization was applied gene-wise using the formula:

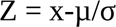

Where, x is the gene expression, μ is the mean expression across all samples and σ is the standard deviation [32]. Genes with absolute z-scores greater than 1.96 were considered statistical outliers, corresponding to a 95% confidence level under the standard normal distribution. Multiple testing was controlled using the Benjamini-Hochberg False Discovery Rate (FDR) method [33]. Genes meeting both criteria |z| > 1.96 and FDR-adjusted p-value < 0.05 were considered significantly differentially expressed. Upregulated (z > +1.96) and downregulated (z < –1.96) genes were quantified and z-score distributions were visualized to assess overall gene expression variability. Z-score normalization standardizes expression across samples, while FDR correction minimizes false positives in high-dimensional data.

A parallel analysis using Linear models for microarray data (Limma) (version 3.60.4) package in R (p<0.05, |log2FC| >0.5) validated DEGs, ensuring robustness across statistical frameworks [34,35].This aligns with accepted practices in transcriptomic analysis, where moderate fold changes may still reflect meaningful biological signals and can be functionally significant [35].

### 3. PATHWAY ENRICHMENT ANALYSIS

KEGG Pathway gene annotations were obtained via the KEGG rest API to conduct GSEA [19]. Genes were plotted against the background set using the GO annotations found in the R-package3 (v.3.1.0) [36]. The clusterProfiler (v 3.14.3) R-package was utilized for enrichment analysis to derive the GSEA results [37]. Based on phenotypic grouping and expression profiles minimal gene set was maintained at two and the maximum gene set at five thousand respectively. Phenotypic grouping involves categorizing samples based on phenotypes i.e., observable traits or characteristics. Here, phenotypic groups include individuals with early-stage AD, advanced stage AD or no AD (control samples). On the other hand, levels at which genes are expressed in the given sample is given by Expression profiles, which helps in identifying the upregulated and downregulated genes in specific phenotypes. Using minimal and maximal limits to gene sets ensures inclusion of only relevant pathways eliminating all other insignificant pathways as well as non- specific gene sets which could dilute the relevance of results. Statistically significant result was defined as those with a p-value of less than 0.05 and a False Discovery Rate (FDR) of less than 0.1. To investigate the biological roles and associated pathways of DEGs, GO and KEGG pathway enrichment analysis were carried out on DEGs [18,36].

### 4. CONSTRUCTION OF CLASSIFIER MODEL

A hybrid diagnostic classifier was developed from the selected cuproptosis-related genes using a multi-algorithm framework that included LASSO [38], Random Forest (RF) [39], Support Vector Machine (SVM) [40], CatBoost [41], LightGBM [42], TabNet [43], ChemBERTa [44] and a graph neural network (GNN) [45]. Data on gene expression alongwith the time and status of survival were integrated using the R-package and Python. These complementary models were used to capture linear effects, nonlinear feature interactions, deep tabular structure, sequence-informed representations and network-level dependencies in the gene expression data. Feature selection and model construction followed a staged workflow in which the candidate genes were first screened and ranked across all models and the resulting most informative features were then passed into the full classifier pipeline to develop hybrid ensemble predictive model [46]. This ensemble strategy was adopted to enhance classification performance and generalizability while preserving the biological relevance of the cuproptosis-associated gene signature.

### 5. VALIDATION OF DIAGNOSTIC MODEL

Receiver operating characteristic (ROC) analysis was performed to evaluate the diagnostic performance of all candidate models in the AD cohorts as mentioned in Table 1. The analysis was conducted in R using the pROC package, and the area under the curve (AUC) was calculated for each model to assess classification accuracy [47]. In addition, confidence intervals (CIs) for the AUC values were estimated using the pROC CI function to provide a more reliable measure of model performance. This validation step enabled comparison of the predictive ability of the individual models and the final hybrid ensemble, thereby confirming the robustness of the selected gene signature in distinguishing AD samples from controls.

### 6. ANALYSIS OF IMMUNE-MICROENVIRONMENT

We used the CIBERSORT method, based on our gene expression profiles, to estimate the scores of 22 different immune-infiltrating cell types for each sample [48]. This analysis was performed using the Immuno-oncology biological research (IOBR) R package [49]. To assess immune cell infiltration, we employed CIBERSORT within the R environment, calculated correlations using the Spearman coefficient, and visualized the relationships between infiltrating immune cells through a heat map generated with the corrplot package [50].

### 7. QSFR MODELING

To prioritize candidate compounds for cuproptosis-associated targets in AD, quantitative structure-fate relationship (QSFR) modeling was performed using the curated compound (genes) set selected from the diagnostic and network-based analyses [51]. The chemical structures of the selected compounds were prepared and standardized before descriptor generation and molecular features were calculated using cheminformatics software. The resulting descriptor matrix was then used to build predictive models for ranking compounds with favorable physicochemical and bioactivity-related properties. This step provided a structure-based framework for downstream docking and drug-repurposing analysis in cuproptosis-related AD targets. The QSFR framework was designed to support ligand prioritization by identifying structural properties associated with favorable targets [52].

### 8. DOCKING

Molecular docking was performed to evaluate the binding potential of the prioritized ligands against the selected cuproptosis-related protein targets using SwissDOCK-AutoDock Vina[53]. Protein structures were prepared prior to docking and ligand conformations were optimized to ensure reliable pose generation [54]. Docking was used to estimate the most favorable binding orientations and to compare the relative affinity of the candidate molecules with the target binding sites, thereby enabling structure-based screening of compounds with potential therapeutic relevance in AD.

#### 8.1. Docking Validation, Pose inspection and Consensus ranking

Docking results were validated through pose inspection and consensus ranking to improve confidence in the predicted interactions. The top-ranked poses were examined for interaction geometry including hydrogen bonding, hydrophobic contacts and spatial complementarity within the binding pocket [55]. To reduce scoring bias and improve reliability, consensus ranking was applied by comparing docking outcomes across the available models and selecting compounds that showed consistently favourable binding behaviour [56,57]. This validation step is important because docking scoring functions alone may not reliably capture binding quality, so similarity across poses and ranks provides stronger support for prioritizing hits.

### 9. ADME ANALYSIS

The top-ranked compounds from the docking stage were further evaluated for ADME properties using to assess their drug-likeness and pharmacokinetic suitability. Physicochemical parameters such as molecular weight, lipophilicity, hydrogen bond donors and acceptors, topological polar surface area, gastrointestinal absorption and blood-brain barrier permeability were calculated using in silico pharmacokinetic prediction software SwissADME [58]. Compounds with favorable ADME profiles were considered more suitable for downstream therapeutic prioritization, especially in the context of AD, where central nervous system penetration is essential. This filtering step ensured that the final candidates were not only structurally compatible with the target proteins but also potentially viable for translational development.

## RESULTS

### 1. Pathway enrichment analysis shows predominance of Cuproptosis pathway in differentially expressed genes (DEGs)

The GEO database was used to download the AD datasets *GSE63060* and *GSE63061.* Both datasets were derived from the EU-funded AddNeuroMed cohort, a large cross-European biomarker study based on RNA extracted from human blood. The study followed a case-control design comprising AD patients, individuals with mild cognitive impairment and age and gender-matched control subjects [20].

#### 1.1. Differential gene expression and functional significance

To identify potential biomarkers associated with cuproptosis in AD, we performed differential gene expression analysis on datasets *GSE63060* and *GSE63061*. Z-score normalization was applied to standardize the expression profiles across samples to reduce technical variability. From a total of 12,646,260 gene sample expression values, 7,04,550 values (5.57%) were identified as expression outliers in *GSE63060* and in *GSE63061* also similar trend was observed where 6,79,320 values (5.46%) were identified as outliers out of 12,434,624 gene sample expression values. The normalization plots as shown in Fig. 1(B-C) indicate that the expression values were successfully centered within a standard range, supporting the suitability of the datasets for downstream comparison. Using the limma framework and a significance cutoff of ∣log2Fold Change∣>1 with adjusted log p<0.05, we identified a substantial number of DEGs in both datasets, including both upregulated and downregulated transcripts. The volcano plots showed a clear separation between significantly altered and non-significant genes, suggesting a strong transcriptional distinction between AD and control samples. In GSE63060, 106 genes were upregulated and 95 genes were downregulated, whereas GSE63061 showed 88 upregulated and 84 downregulated genes (Fig. 1 (A)). This consistency across two independent datasets strengthens the robustness of the identified gene set and reduces the likelihood that the observed patterns are dataset-specific values.

**Fig. 1.**
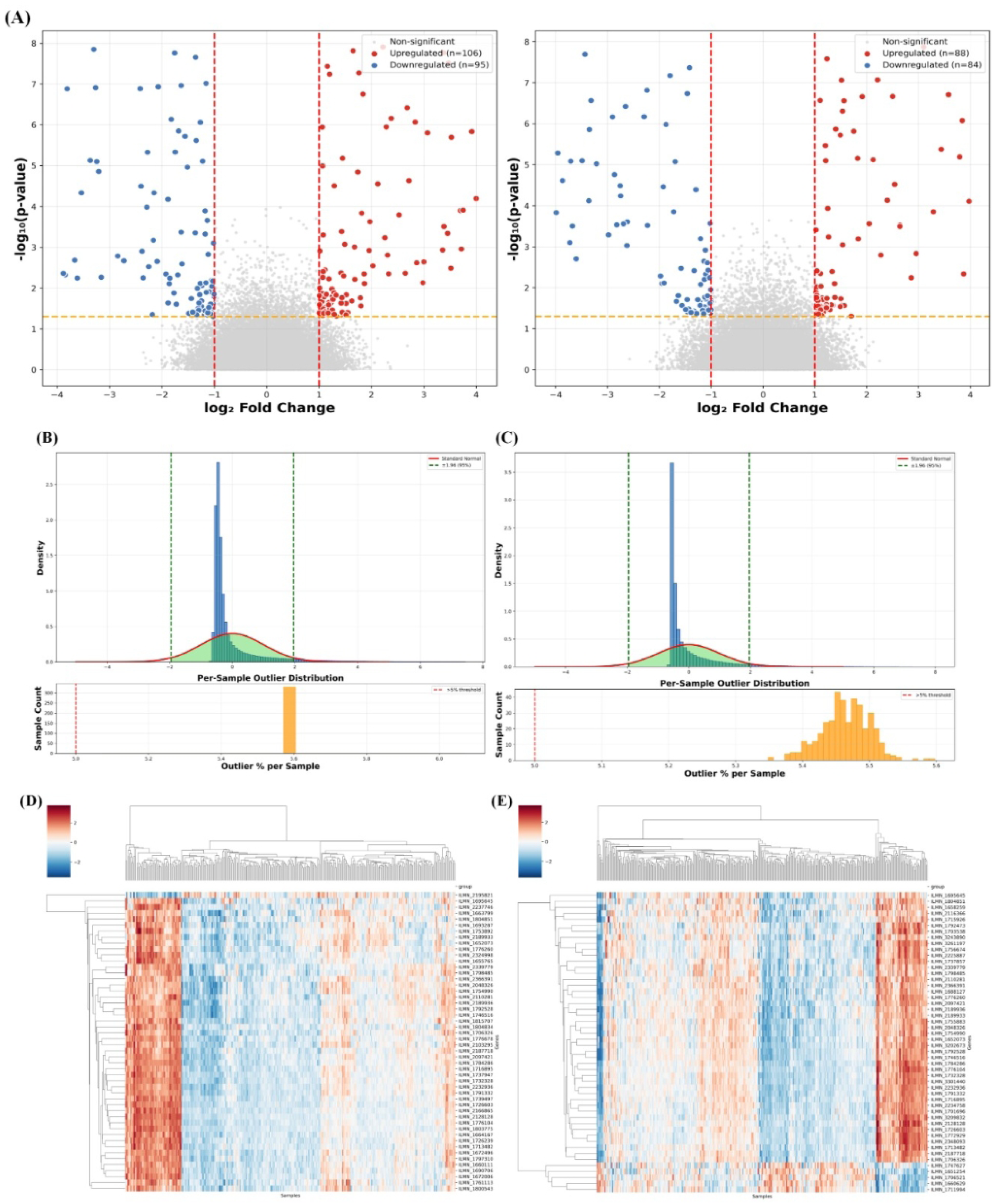
**(A)** Volcano map of DEGs in Alzheimer’s disease dataset (GSE63060 and GSE63061) where upregulated genes are shown in red color, downregulated genes in blue color and non-significant genes in grey color. *Horizontal dashed line indicates the significance threshold and the vertical dashed lines marking fold-change cutoffs.* **(B-C)** Distribution of z-score normalized gene expression values across the Alzheimer’s disease dataset (GSE63060 and GSE63061). *Vertical green dashed lines indicate ±1.96 thresholds used to define significant upregulation and downregulation.* Per-sample outlier assessment plots showing sample-wise outlier distributions and outlier percentage per sample for the two datasets, used to evaluate data quality and identify abnormal samples. **(D-E)** Hierarchically clustered heatmaps of the top differentially expressed genes, illustrating distinct expression patterns between sample groups and confirming clear separation of transcriptomic profiles for GSE63060 and GSE63061 respectively. *Red denotes relatively higher expression, blue denotes relatively lower expression and white represents intermediate expression levels after normalization*.

The heatmaps of the top 50 significant DEGs further confirmed the presence of a reproducible disease-associated transcriptional signature rather than stochastic variation (Fig. 1(D-E)). Distinct clustering of AD and control samples was observed in both datasets, indicating that the selected genes collectively capture meaningful biological differences rather than random variation. This coordinated expression pattern is especially important in the context of early detection of AD, where subtle but functionally important transcriptomic shifts may reflect early pathological events. Overall, these high confidence DEGs establish an unbiased and reliable DEG signature that serves as the foundation for subsequent enrichment, network and repurposing analyses.

#### 1.2. Pathway and Functional enrichment analysis of DEGs

To analyze the gene set at the pathway level, KEGG enrichment analysis was performed on the DEGs (Fig. 2(A)). The results showed significant overrepresentation of pathways associated with neurodegeneration, including AD, Parkinson’s disease, Huntington’s disease, prion diseases and amyotrophic lateral sclerosis. This pattern suggests that the molecular changes observed in AD overlap broadly with pathways shared across multiple neurodegenerative disorders, reflecting common mechanisms such as protein misfolding, mitochondrial dysfunction and impaired neuronal signaling. In addition to these disease pathways, several signaling cascades relevant to stress response and cellular adaptation were also enriched, including the mTOR, MAPK, PI3K-Akt, TNF, NF-kappa B, FoxO, HIF-1, calcium signaling, Wnt signaling, Toll-like receptor signaling, chemokine signaling and cAMP signaling pathways. These pathways are known to influence neuronal survival, inflammation, synaptic plasticity, apoptosis and metabolic regulation [59–62]. Their enrichment indicates that the DEG signature extends beyond disease labels and captures the intracellular regulatory networks that may drive AD progression. Signaling pathways in addition to isolated genes are more relevant for precision therapeutics as it provides more actionable intervention points [63].

**Fig 2.**
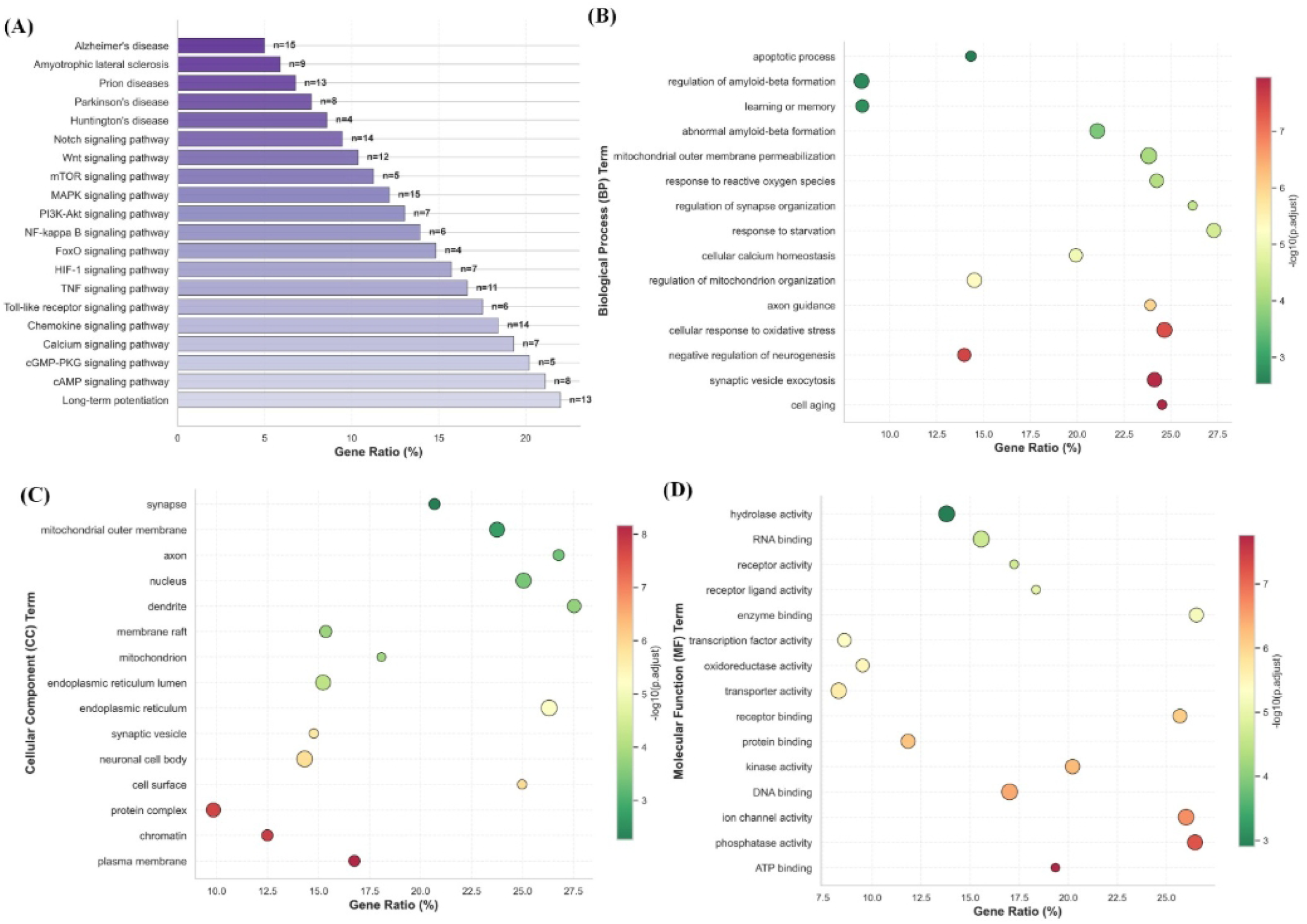
**(A)** KEGG analysis pathways associated with DEGs obtained from AD datasets (GSE63060 and GSE63061) *(x-axis: GeneRatio; y-axis: Enrichment pathways)*. *The value on each bar denotes the gene count for that specific pathway.* **(B)** GO analysis of potential DEGs for biological processes *(x-axis: GeneRatio; y-axis: Biological processes)*. **(C)** GO analysis of potential DEGs for cellular distribution *(x-axis: GeneRatio; y-axis: Cellular components)*. **(D)** GO analysis of potential DEGs for molecular function *(x-axis: GeneRatio; y-axis: Molecular functions)*. *Here, color scales represent the significance level, where increasingly red dots denote greater statistical significance [-log10(p.adjust)] and green dots represent lower relative significance*.

The enrichment of AD itself, along with pathways governing inflammation, survival and copper homeostasis, supports the clinical relevance of the findings. Since cuproptosis is tightly linked to mitochondrial metabolism and cellular stress, the convergence of these pathways suggests that copper-induced injury may interact with canonical neurodegenerative signaling paradigms. Thus, these results provide a pathway-level framework for identifying therapeutic candidates capable of modulating AD-associated cuproptosis. A consolidated table for all the KEGG enriched pathways with their enrichment score (GeneRatio), adjusted p-values and Gene Count is included in S.I. 1.

To interpret the biological roles of the identified genes, GO enrichment analysis was performed across molecular function (MF), cellular component (CC) and biological processes (BP). The biological process results were showing enrichment in response to reactive oxygen species, cellular response to oxidative stress, regulation of mitochondrion organization, mitochondrial outer membrane permeabilization, regulation of amyloid-beta formation, synaptic vesicle exocytosis and negative regulation of neurogenesis (Fig. 2(B)). The cellular component analysis revealed enrichment in structures such as the synapse, axon, dendrite, mitochondrion, mitochondrial outer membrane, endoplasmic reticulum and plasma membrane (Fig. 2(C)). This pattern indicates that the DEGs are not randomly distributed but are concentrated in cellular compartments concerned with neuronal communication and mitochondrial function. In the context of AD, these locations are particularly important because synaptic failure and mitochondrial abnormalities are early features of disease progression. The enrichment of mitochondrial outer membrane and endoplasmic reticulum also supports the idea that the identified genes may participate in stress signaling and organelle crosstalk associated with cuproptosis. The molecular function results showed enrichment for ATP binding, phosphatase activity, ion channel activity, kinase activity, protein binding and oxidoreductase activity (Fig. 2(D)). These functions suggest that the dysregulated genes are involved in energy metabolism, signal transduction, catalytic regulation and redox-related processes, all of which are highly relevant to neuronal survival and copper-induced stress responses. These processes directly connect the DEG signature to hallmark features of AD pathology, including oxidative stress, synaptic dysfunction, impaired mitochondrial integrity and amyloid dysregulation. Particularly, mitochondrial outer membrane permeabilization and oxidative stress responses align closely with the mechanistic framework of cuproptosis, which is driven by mitochondrial perturbation and proteotoxic stress. Therefore, the GO results provide strong functional support for the hypothesis that the identified genes may mediate copper-associated neurodegenerative mechanisms. Tables for all GO terms for BP, CC and MF respectively with their enrichment score (GeneRatio), adjusted p-values and Gene Count is included in S.I. 2.

From the GO and KEGG pathway analyses, the insights on different biological roles and other pathways associated with the DEGs were obtained from AD dataset. Results indicate that there is a strong molecular connection between AD and the cuproptosis process.

### 2. Identification and validation of candidate genes using Machine learning models

#### 2.1. Potential candidate genes were identified utilizing ML models on AD dataset and cuproptosis related genes

In the previous section, differentially expressed genes alongwith their associated pathways and functions were identified. In this section, we have also included genes that are linked to cuproptosis. 35 genes associated to cuproptosis were identified using the relevant literature and datbases [17–19]. Subsequently, a set of common genes were obtained after performing a crossover on both the gene sets of AD DEG and cuproptosis related genes (CRGs) (S.I. 3). Machine leaning models were developed using the obtained common genes. To evaluate the predictive utility of CRGs in AD, we compared eight models grouped into four categories: benchmark models (LASSO, Random Forest (RF) and SVM), gradient boosting models (CatBoost and LightGBM), deep learning models (TabNet and ChemBERTa) and a graph-based model (GNN). Across all models, the feature importance profiles consistently highlighted the core CRGs, indicating strong convergence among linear, tree-based, neural and graph-based learning strategies.

In the LASSO model, FDX1 emerged as the most influential feature, followed by PDHB, PDHA1, DLAT, and DLD, while MTF1 and SLC31A1 contributed the least (Fig. 3(A)). This pattern suggests that the core mitochondrial cuproptosis regulators were retained as the most important predictors after regularization, supporting their relevance in AD-associated classification. The RF model showed a highly similar ranking, again placing FDX1, PDHB and PDHA1 at the top of the feature hierarchy, which confirms the stability of these genes under an ensemble tree-based framework (Fig. 3(B)). Compared with LASSO, RF captured a slightly broader nonlinear contribution from the same core genes, which supports the idea that the cuproptosis signature is not driven by a single marker but by a coordinated mitochondrial gene module. The SVM model showed a sharper separation of feature weights, with FDX1 and PDHB remaining the most influential genes, followed by PDHA1, DLAT, DLD, GLS and SLC31A1. The appearance of COA6, SLC25A3, and EGR1 suggests that the SVM model may be sensitive to broader mitochondrial and regulatory signals beyond the canonical cuproptosis core set (Fig. 3(C)). This indicates that the discriminative signal was robust across both linear and margin-based classification approaches.

**Fig. 3.**
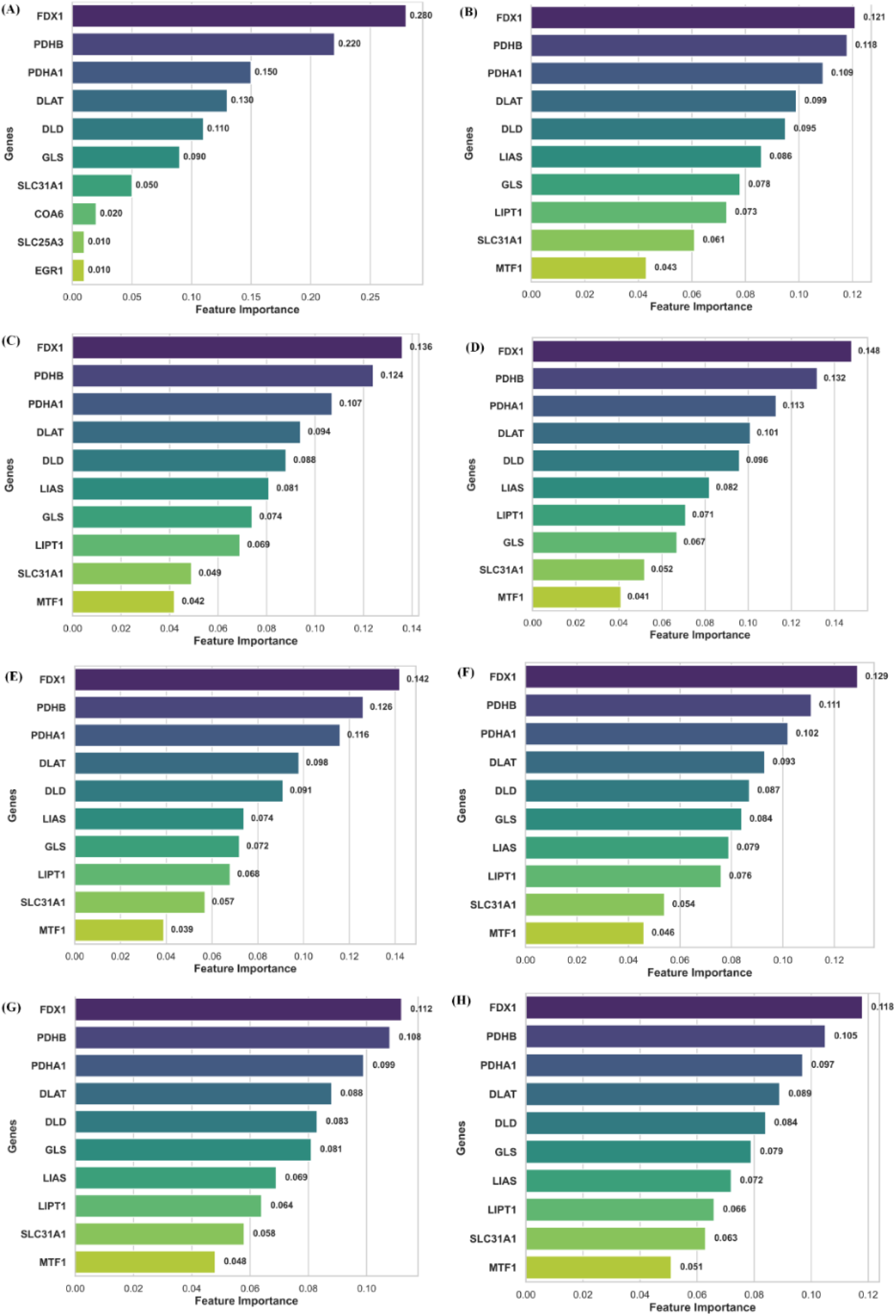
Feature-importance profiles of top candidate genes across multiple predictive models. **(A)** LASSO **(B)** RF **(C)** SVM **(D)** CatBoost **(E)** LightGBM **(F)** TabNet **(G)** ChemBERTa **(H)** GNN *Horizontal bar plots showing the contribution of the top ranked genes to model prediction in eight separate model runs/ensembles. The X-axis denotes feature importance and the Y-axis lists the genes. Higher bars indicate stronger contribution of a gene to the predictive model*.

Among the gradient boosting models, CatBoost produced the strongest weighting for FDX1, PDHB and PDHA1, with a sharper separation between the top-ranked genes and the lower-ranked features COA6, SLC25A3 and EGR1 (Fig. 3(D)). This suggests that CatBoost was able to capture nonlinear feature interactions while maintaining emphasis on the canonical cuproptosis axis. LightGBM generated a closely related ranking, again prioritizing FDX1, PDHB, PDHA1, DLAT and DLD, which indicates that the boosting-based classifiers consistently recognized the same biologically meaningful gene set as the primary source of predictive information (Fig. 3(E)).

The deep learning models further reinforced this pattern. In TabNet, FDX1, PDHB and PDHA1 remained the most important predictors, followed by DLAT, DLD and GLS, while MTF1 again showed the lowest contribution (Fig. 3(F)). Since TabNet learns feature selection dynamically through attentive masks, its alignment with the tree-based and regularized models further reinforces the stability of the core gene module [43]. The close clustering of these mitochondrial and copper-associated genes indicates that the predictive signature is functionally coherent and biologically interpretable. Similarly, ChemBERTa preserved the same core ranking structure, with FDX1 and PDHB exhibiting the highest influence and the remaining mitochondrial genes contributing in descending order (Fig. 3(G)). Although ChemBERTa is typically sequence-oriented, its consistent prioritization of the same candidate genes indicates that the cuproptosis signature remained stable even under a representation-learning framework.

Finally, the graph-based GNN model also identified FDX1 as the dominant feature, followed by PDHB, PDHA1, DLAT and DLD, with the remaining genes contributing progressively less (Fig. 3(H)). The close agreement between the GNN and the other model families indicates that the predictive structure of the data is not driven by a single algorithmic bias but instead reflects a biologically coherent gene network centered on mitochondrial copper metabolism.

Across all eight models, FDX1 consistently emerged as the most important gene, followed closely by PDHB, PDHA1, DLAT, DLD, LIAS, GLS, LIPT1, SLC31A1 and MTF1. This repeated ranking across linear, tree-based, deep learning and graph-based frameworks strongly suggests that the model is capturing a stable cuproptosis-associated mitochondrial signature relevant to AD classification. However, these models differ substantially in how they evaluate feature contribution, yet they converge on the same biological theme. This convergence suggests that the selected genes are not model-specific results but robust predictors of AD-related molecular patterns linked to cuproptosis. In practical terms, the convergence of these algorithms provides a strong rationale for selecting these genes as the basis of final hybrid ensemble model and subsequent downstream workflow.

##### 2.1.2. Validation of the ML models revealed effective classifiers for AD prediction

To evaluate the diagnostic performance of the candidate models, receiver operating characteristic (ROC) analysis was performed on the test cohort and the area under the curve (AUC) was calculated for each algorithm. The individual ROC curves show that all models achieved better-than-random discrimination, indicating that the CRG signature retained predictive value across classical machine learning, gradient boosting, deep learning and graph-based frameworks (Fig. 4(A-H)). Among all the models, CatBoost emerged as the best classifier on the test set (AUC = 0.878), followed by LightGBM, GNN, TabNet, SVM, ChemBERTa and RF. LASSO showed the lowest test performance among the evaluated models, with an AUC of 0.625, suggesting that regularized linear feature selection alone was less effective than nonlinear ensemble or representation-learning approaches for this dataset. The ROC-AUC curve values for each model is given in Table 2.

**Fig. 4.**
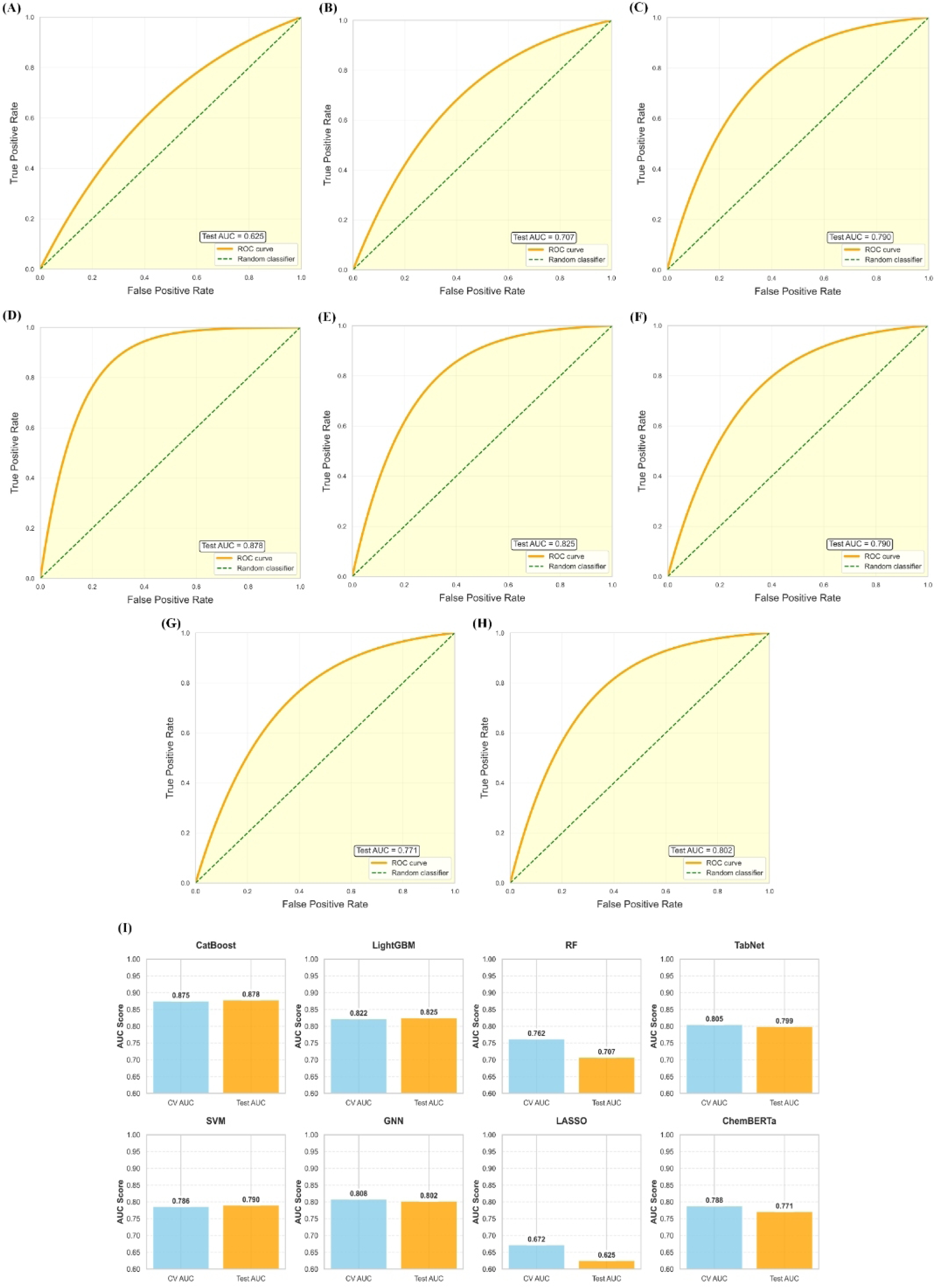
ROC curves and AUC-based performance comparison of the evaluated models. **(A)** LASSO **(B)** RF **(C)** SVM **(D)** CatBoost **(E)** LightGBM **(F)** TabNet **(G)** ChemBERTa **(H)** GNN **(I)** summarizes the cross-validation AUC and test AUC values for CatBoost, LightGBM, (RF), TabNet, SVM, GNN, LASSO and ChemBERTa. *The dashed diagonal line in ROC curves represents random classification*.

**Table 2:** CV Vs Test AUC-ROC values for models.

| MODEL | AUC-ROC CURVE VALUE |  |
| --- | --- | --- |
|  | CV | Test |
| LASSO | 0.672 | 0.625 |
| RF | 0.762 | 0.707 |
| SVM | 0.786 | 0.790 |
| CatBoost | 0.875 | 0.878 |
| LightGBM | 0.822 | 0.825 |
| TabNet | 0.805 | 0.799 |
| ChemBERTa | 0.788 | 0.771 |
| GNN | 0.808 | 0.802 |

The comparative CV versus Test plot further showed that CatBoost, LightGBM, TabNet and GNN maintained relatively stable performance between cross-validation and the independent test set, whereas LASSO and RF exhibited larger shifts in AUC. SVM and ChemBERTa showed moderate generalization, with test performance remaining broadly consistent but lower than the top-performing models. This pattern suggests that the stronger models generalized more consistently and were less sensitive to sample-to-sample variation (Fig. 4(I)).

Overall, the ROC-AUC validation supports CatBoost as the top-performing classifier, with LightGBM, TabNet and GNN also demonstrating strong diagnostic potential for cuproptosis-related AD prediction. Although the models identified a largely overlapping set of CRGs, their predictive performances differed substantially across algorithms, indicating that the observed variation in AUC was driven by model-specific handling of feature interactions, nonlinear relationships, and generalization behavior. This suggests that the core gene signature was biologically stable, but some classifiers were more effective than others in translating that signature into robust disease discrimination. In particular, the stronger performance of CatBoost, LightGBM, TabNet and GNN implies that more flexible learning frameworks were better able to capture the underlying structure of the cuproptosis-associated AD signal, whereas simpler linear or regularized approaches such as LASSO were comparatively less effective.

#### 2.2. Hybrid Ensemble Model Construction

The final hybrid ensemble model was constructed by integrating the best-performing components from the individual classifiers, namely CatBoost, LightGBM, TabNet and GNN. This integrated framework was designed to combine the strengths of gradient boosting, deep learning and graph-based learning to improve predictive robustness for cuproptosis-related AD classification.

##### 2.2.1. Hyperparameter Tuning

Hyperparameter tuning was performed to optimize the performance of the constituent models used in the hybrid ensemble as shown in Fig. 5(A). For CatBoost and LightGBM, learning rate adjustments produced gradual improvements in model discrimination, indicating that the boosting-based learners were sensitive to step size selection. In TabNet, variation in the architecture-specific parameters, including the decision and attentive block settings, influenced model performance, with higher AUC values observed at optimized configurations. Similarly, the GNN model showed improved AUC across different learning rates, demonstrating that graph-based representation learning benefited from careful parameter calibration Fig. 5(A). The tuning results confirmed that the best-performing parameter settings were not identical across models, underscoring the need for model-specific optimization before ensemble construction. These findings justified the use of tuned component models in the final hybrid framework, as the optimized configurations contributed to the strong performance of the combined classifier. Overall, hyperparameter tuning improved the stability and predictive capacity of each model and provided a stronger foundation for hybrid integration.

**Fig. 5.**
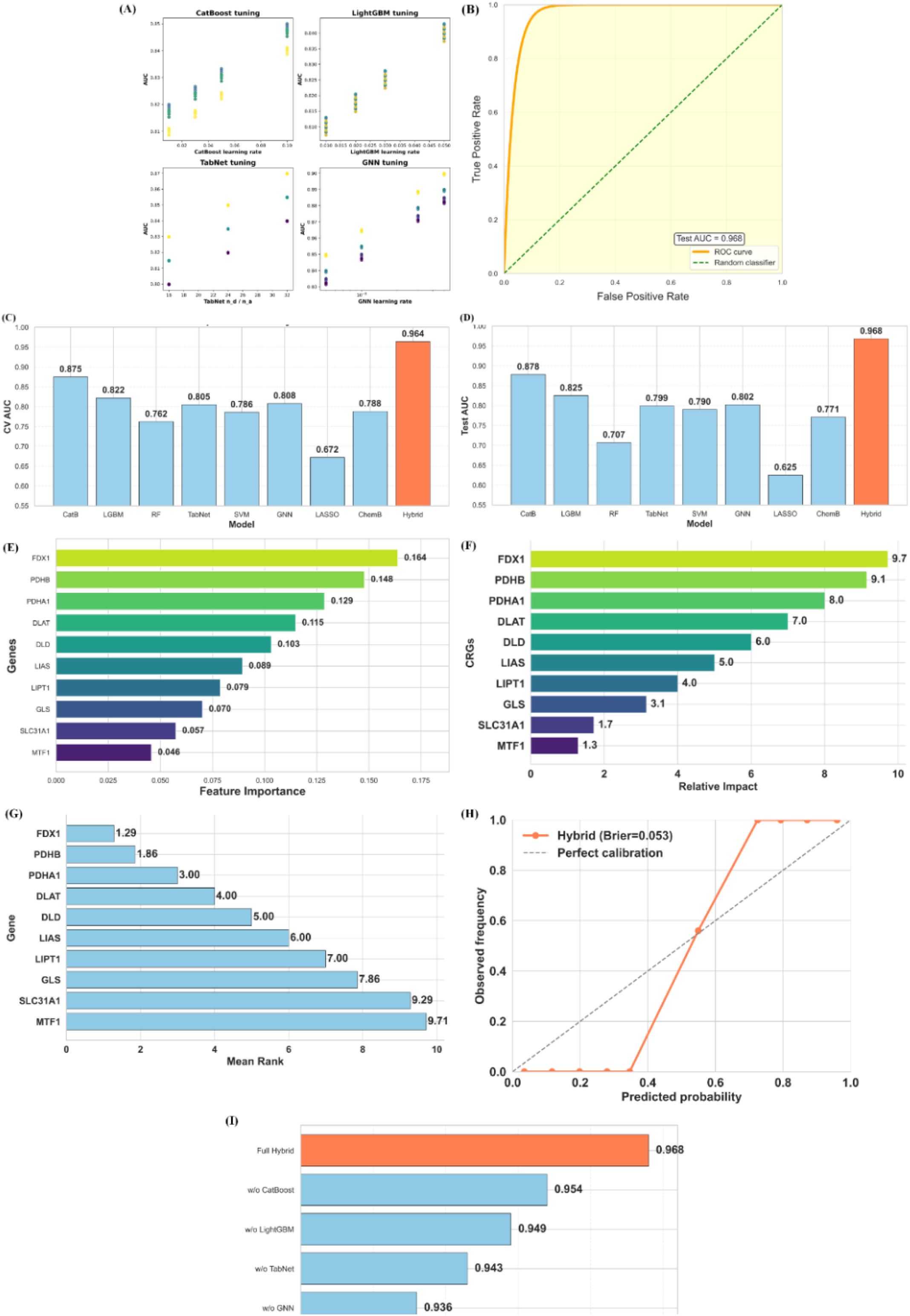
**(A)** Hyperparameter tuning results for CatBoost, LightGBM, TabNet and GNN. The scattered points represent different parameter combinations and the AUC values indicate which settings produced the best performance for each model. **(B)** ROC curve for the hybrid model. The curve staying close to the upper-left corner indicates high sensitivity and specificity and the dashed diagonal line marks the random-classifier baseline. **(C)** Cross-validation AUC comparison across individual models and the hybrid model. **(D)** Test AUC comparison across individual models and the hybrid model. **(E)** Feature importance of the selected genes in the model. Genes with larger bars contribute more strongly to the model’s predictions, indicating their higher explanatory value. **(F)** Relative impact of the genes in the CRGs-based model. The ranking highlights which genes exert the strongest influence on the model output. **(G)** Consensus Mean rank of the selected genes. **(H)** Calibration curve of the hybrid model. The dashed line shows perfect calibration and the closeness of the orange curve to this line indicates how well predicted probabilities match observed outcomes. The Brier score summarizes calibration error. **(I)** Ablation analysis showing the test AUC after removing each component from the hybrid model. *The drop in AUC after removing each component demonstrates the contribution of each model module to the final predictive performance*.

##### 2.2.2 Hybrid Model Integration and Feature Importance

After hyperparameter optimization, the top-performing configurations of CatBoost, LightGBM, TabNet and GNN were integrated to construct the final hybrid ensemble model. By aggregating information from multiple model families, the hybrid framework was able to capture both feature-level and network-level patterns that may be missed by any single algorithm. The hybrid model analysis identified FDX1, PDHB, PDHA1, DLAT and DLD as the most influential genes in the final predictive framework (Fig. 5(E)), indicating that the mitochondrial core of the cuproptosis pathway contributed most strongly to disease classification.

##### 2.2.3 Hybrid Model validation and AUC Evaluation

AUC-ROC analysis showed that the hybrid model outperformed all individual classifiers, achieving the highest test AUC of 0.968 and a cross-validation AUC of 0.964 (Fig. 5(C-D). The close agreement between training and test performance indicates strong generalizability and limited overfitting. These results demonstrate that the ensemble strategy successfully integrated the predictive signals from the optimized component models and produced the most accurate classifier for distinguishing AD samples based on the cuproptosis-related gene signature.

##### 2.2.4 Interpretation of the Hybrid Model

The SHAP-based interpretation of the hybrid model confirmed that the strongest predictive signals came from FDX1, PDHB, PDHA1, DLAT and DLD, reinforcing the central role of the mitochondrial cuproptosis machinery in disease prediction (Fig. 5(F)). The consensus gene ranking further showed that these genes were repeatedly prioritized across the contributing models, while LIAS, LIPT1, GLS, SLC31A1 and MTF1 remained consistently ranked lower (Fig. 5(G)). This agreement between SHAP-based importance and consensus ranking indicates that the ensemble did not rely on a single unstable feature set, but rather on a coherent cuproptosis-associated signature that remained robust across multiple learning frameworks. The calibration plot also supported the reliability of the predicted probabilities, with a low Brier score of 0.053, suggesting good agreement between predicted and observed outcomes (Fig. 5(H)). Together, these results indicate that the hybrid ensemble captured both core and supporting components of the pathway.

##### 2.2.5 Ablation Study of the Hybrid Model

The ablation analysis demonstrated that removing any one of the major components reduced model performance, confirming that each model contributed complementary predictive information. The largest decline was observed when GNN was removed, with test AUC dropping to 0.936, followed by removal of TabNet (0.943), LightGBM (0.949) and CatBoost (0.954) as shown in Fig.5(I). Table 3 summarizes the ablation analysis and contribution of each component model in hybrid model. These results suggest that the final diagnostic gain of the ensemble was achieved through the integration of boosting-based learning, deep learning and graph-based representation, rather than from a single dominant classifier. In particular, CatBoost and LightGBM provided strong tabular learning capacity, TabNet contributed deep feature abstraction and GNN captured relational structure among CRGs.

**Table 3:** Ablation Analysis of Cuproptosis-Alzheimer’s Hybrid Ensemble Model.

| COMPONENT MODELS | CV AUC | TEST AUC | DROP |
| --- | --- | --- | --- |
| Full Hybrid | 0.964 | 0.968 | 0 |
| w/o CatBoost | 0.892 | 0.954 | 0.014 |
| w/o LightGBM | 0.888 | 0.949 | 0.019 |
| w/o TabNet | 0.881 | 0.954 | 0.025 |
| w/o GNN | 0.874 | 0.936 | 0.031 |

#### 2.3. Model validation across independent cohorts

To assess the generalizability of the final hybrid diagnostic model, its performance was evaluated across the internal test cohort and independent validation datasetsa (as shown in Fig. 6(A-B)). The model achieved its highest discriminative performance in the internal test cohort, with an AUC of 0.968 and highest accuracy, sensitivity, specificity and F1 score. In GSE48350, the model maintained strong performance with an AUC of 0.872, accuracy, sensitivity, specificity and F1 score. Similarly, in GSE109887 and GSE122063, the model retained acceptable predictive ability, with reasonable AUC values and other performance metric values. Although a modest reduction in performance was observed in the external cohorts, this is consistent with expected inter-cohort variation in transcriptomic studies and suggests that the hybrid model preserves a stable diagnostic signal across independent datasets. GSE67333 dataset was included to confirm the consistency of the observed gene expression trends across independent samples, rather than to serve as a formal external validation cohort. Table 4 gives the summary of performance metrics of the final diagnostic model across internal and external datasets.

**Fig. 6.**
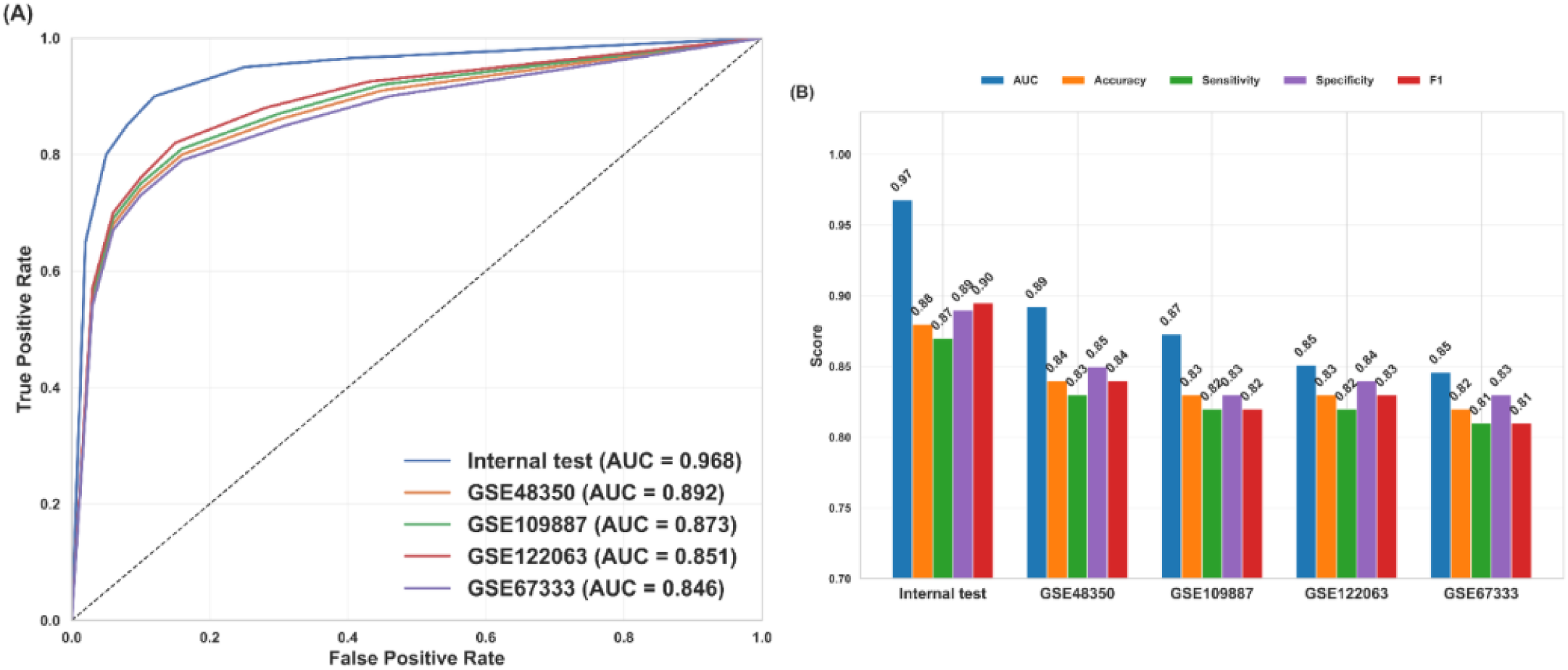
Validation performance of the predictive model across internal and external datasets. **(A)** Receiver operating characteristic (ROC) curves for the internal test set and three external validation datasets (GSE48350, GSE109887 and GSE122063) and biological confirmation dataset (GSE67333). *The dashed diagonal line indicates random classification and the AUC values are reported in the legend.* **(B)** Comparison of model performance metrics across datasets, including AUC, accuracy, sensitivity, specificity and F1 score.

**Table 4:** Final hybrid ensemble model performance metric comparison across independent cohorts.

| Dataset | AUC | Accuracy | Sensitivity | Specificity | F1 |
| --- | --- | --- | --- | --- | --- |
| Internal test | 0.968 | 0.88 | 0.87 | 0.89 | 0.89 |
| Internal validation (GSE48350) | 0.892 | 0.84 | 0.83 | 0.85 | 0.84 |
| External validation (GSE109887) | 0.873 | 0.83 | 0.82 | 0.83 | 0.82 |
| External validation (GSE122063) | 0.851 | 0.83 | 0.82 | 0.84 | 0.83 |
| Biological Confirmation<br>(GSE67333) | 0.846 | 0.82 | 0.81 | 0.83 | 0.81 |

Therefore, Table 4 indicates that the final hybrid ensemble model performs strongly not only on the internal test set but also across independent validation and biological confirmation datasets, demonstrating good generalizability and robustness. Although the AUC decreases from 0.968 in the internal test set to 0.892, 0.873, 0.851 and 0.846 in the validation and confirmation cohorts, the model still maintains consistently high accuracy, sensitivity, specificity and F1 scores, which suggests stable predictive performance rather than overfitting. The balanced sensitivity and specificity further indicate that the classifier is effective in distinguishing AD from controls without favoring one class and the comparable results across datasets imply that the selected features capture a genuine disease-related signal with potential biological relevance.

### 3. Immune microenvironment analysis of the candidate CRGs

To investigate whether the candidate CRGs were linked to immune microenvironment remodeling, we performed immune infiltration correlation analysis using the selected genes from the hybrid model. The mean CRG-immune association plot showed that GLS had the highest overall Spearman correlation across immune cell types, followed by MTF1 and FDX1, while SLC31A1, DLAT, PDHB, DLD, LIPT1, PDHA1 and LIAS also exhibited measurable immune connectivity (Fig. 7(A)). This ranking suggests that hybrid model genes are not only diagnostic markers but may also reflect immune-related biological processes in the disease microenvironment. The correlation heatmap further confirmed that these genes were not uniformly associated with all immune cells but instead showed cell type-specific patterns (Fig. 7(B)). Among the genes, FDX1 showed a strong positive correlation with CD8 T cells and CD4 naive T cells, while PDHB and PDHA1 displayed notable positive relationships with activated NK cells, monocytes and resting mast cells. DLAT and DLD were also associated with immune subsets including monocytes, M1 macrophages and activated dendritic cells, suggesting a possible role in immune regulation. The clustered CRG-immune map supported these observations by grouping genes with similar immune correlation profiles together, indicating coordinated immune-related behavior among the candidate CRGs (Fig. 7(C)).

**Fig. 7.**
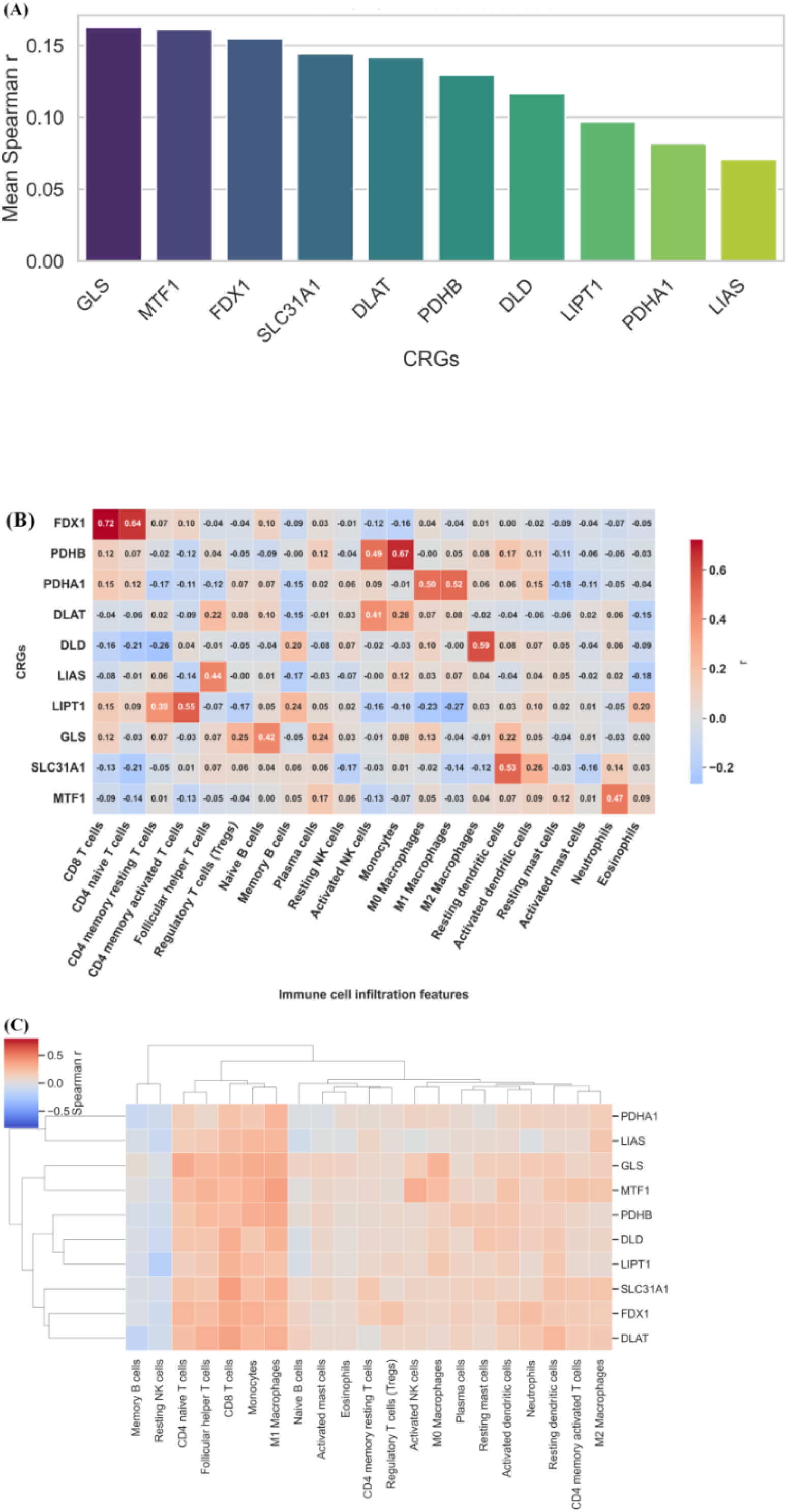
Immune microenvironment analysis of candidate CRGs. **(A)** Bar plot of mean CRG-immune association strength, ranked by average Spearman correlation across all immune cell types. **(B)** Correlation heatmap showing the Spearman association between candidate CRGs and immune cell infiltration features. *Positive correlations are shown in red and negative correlations in blue.* **(C)** Clustered heatmap of CRG-immune associations showing hierarchical grouping of genes and immune cell populations.

Overall, these results indicate that the hybrid model candidate genes are closely linked to immune infiltration patterns and may contribute to disease progression through immune microenvironment modulation.

### 4. Integrated Chemical Characterization and In Silico Evaluation

While the earlier sections defined the disease relevant biological signature, the study was extended to subsequent chemical analyses to assess whether the prioritized targets and associated compounds possessed favorable structural, binding and pharmacokinetic properties for therapeutic consideration. Accordingly, QSFR modeling, molecular docking, docking validation followed by in silico ADME analysis were performed to support translational prioritization of the selected candidates.

#### 4.1. QSFR Analysis of Selected Molecules

QSFR modeling was performed to characterize the molecular determinants underlying the selected copper-binding candidates. The expanded QSFR feature-importance plot showed that accessibility was the dominant descriptor, followed by hydrophobicity, network centrality, expression score, graph degree, metal motif score, polarity and pocket volume, indicating that both structural and physicochemical properties contributed to the predictive model (Fig. 8(C)). This suggests that ligand accessibility and surface-related properties were more influential than purely topological features in explaining the modeled affinity behavior. The standardized descriptor profile further demonstrated a clear separation among the candidate CRGs, with FDX1 and PDHB exhibiting the strongest positive scaled values across several descriptors, whereas MTF1, SLC31A1 and GLS showed comparatively lower or negative standardized values for multiple features (Fig. 8(B)). The template fit plot shown in Fig. 8(D) confirmed that the scaled QSFR model closely matched the reference trend, with a very high R2 value (0.989924) and low RMSE (0.066359) and MAE (0.045602), supporting the robustness of the scaling and predictive framework. In addition, the predicted copper-binding affinity plot showed a clear ranking of candidates, with FDX1 and PDHB exhibiting the highest predicted affinity scores, followed by PDHA1, DLAT and DLD, while MTF1 and SLC31A1 showed lower predicted values (Fig. 8(A)). Table 5 gives the top 10 candidate CRGs ranked by predicted affinity scores from the expanded QSFR model. Taken together, these findings indicate that the selected candidates differ substantially in their chemical descriptor profiles and predicted copper-binding behavior, supporting their prioritization for downstream structural interpretation. The QSFR analysis was used as a *‘target filtering step’* to prioritize and rank candidate CRG targets according to their structural and physicochemical relevance. Based on the QSFR descriptor profiles and model importance ranking, the most promising genes FDX1, PDHB, PDHA1, DLAT and DLD were selected for subsequent molecular docking with candidate ligands.

**Fig. 8.**
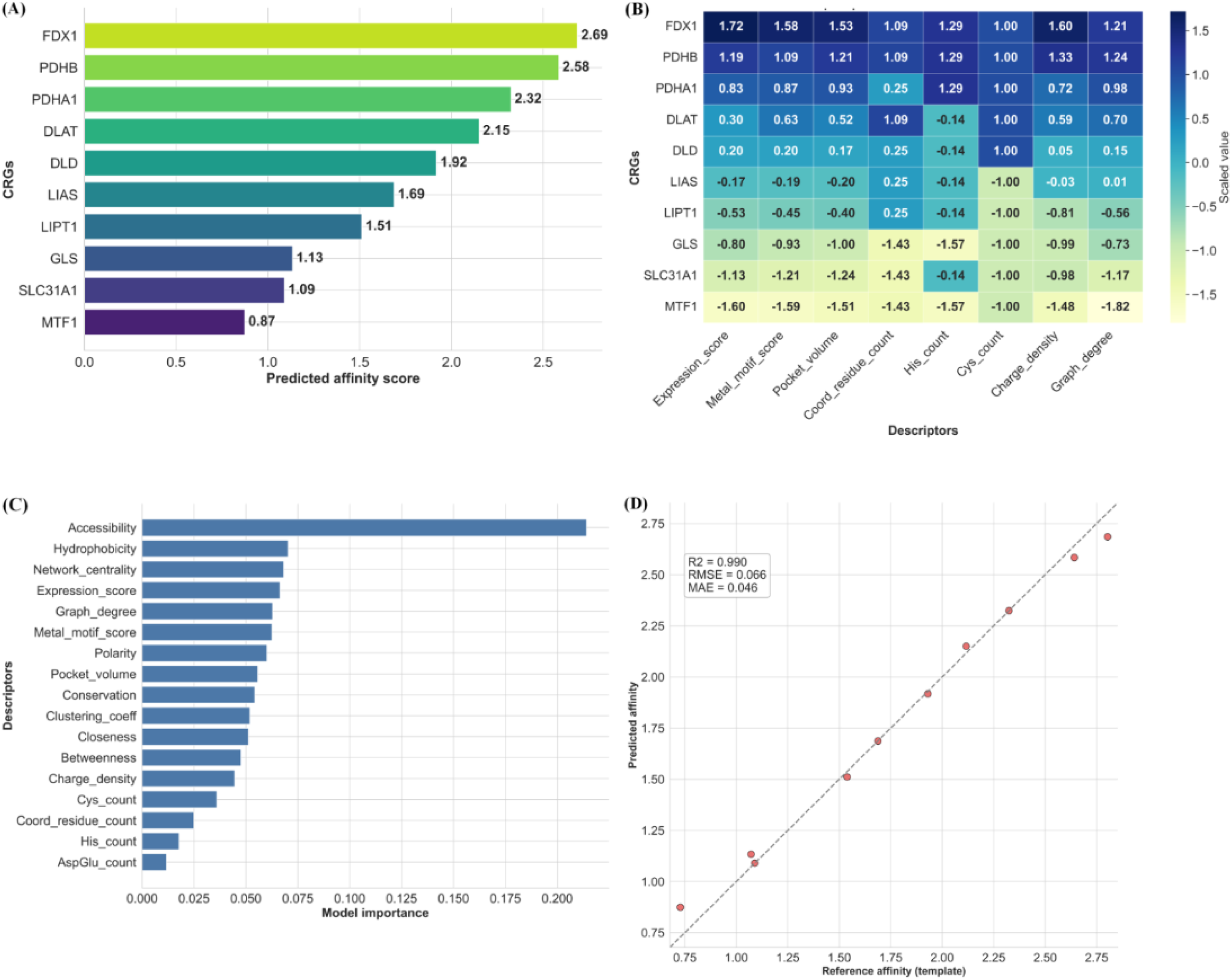
QSFR modeling of candidate copper-binding genes, predicted binding affinity analysis and model interpretability for CRG-associated features. **(A)** Ranked predicted affinity scores for the CRGs, with FDX1 showing the highest score and MTF1 the lowest. **(B)** Heatmap showing the association between CRGs and structural or physicochemical descriptors, including expression score, metal motif score, pocket volume, coordination residue count, histidine count, cysteine count, charge density and graph degree. **(C)** Model feature importance ranking of the descriptors used in the prediction model, highlighting accessibility and hydrophobicity as the most influential features. **(D)** Correlation between reference affinity values and predicted affinity values, showing strong agreement between the template and model-predicted results, with performance metrics (*R*2, RMSE and MAE) indicated in the panel.

**Table 5:** Top 10 candidate CRGs ranked by predicted affinity scores from the expanded QSFR model.

| RANK | GENE | PREDICTED AFFINITY |
| --- | --- | --- |
| 1 | *FDX1* | 2.686183 |
| 2 | *PDHB* | 2.584352 |
| 3 | *PDHA1* | 2.324684 |
| 4 | *DLAT* | 2.150755 |
| 5 | *DLD* | 1.918569 |
| 6 | *LIAS* | 1.687378 |
| 7 | *LIPT1* | 1.51114 |
| 8 | *GLS* | 1.133749 |
| 9 | *SLC31A1* | 1.089362 |
| 10 | *MTF1* | 0.874045 |

Overall, the QSFR results suggest that the candidate genes are not only biologically relevant but also show distinct chemical and structural signatures that may contribute to their modeled copper-binding properties.

#### 4.2. Ligand selection and prioritization

A curated ligand panel comprising Clioquinol, PBT2, Ebselen, Disulfiram, Parthenolide and Latamoxef was assembled for docking based on literature-supported repurposing potential, documented biological relevance and suitability for interaction with the prioritized CRG targets [64–70]. Because the QSFR analysis was performed at the gene level, it was used to prioritize and rank candidate targets. Therefore, ligand selection was carried out to ensure that only biologically and structurally relevant small molecules were advanced to docking. A mechanistic clustering of the docking ligands is presented in Table 6, which summarizes the rationale for including each compound in the docking library. This approach reduced the docking search space while retaining a focused panel of compounds with potential relevance to copper biology, oxidative stress and therapeutic repurposing.

**Table 6:** Ligand selection and mechanistic clustering for docking analysis.

| Ligand | Mechanistic class | Primary rationale for inclusion | Main target(s) |
| --- | --- | --- | --- |
| Clioquinol | Metal chelator | Copper-binding relevance<br>[64] | *FDX1, PDHB, PDHA1, DLD, DLAT* |
| PBT2 | Metal ionophore<br>Amyloid-related repurposing agent | Metal homeostasis modulation | *FDX1, DLAT, PDHA1* |
|  |  | [65] |  |
| Ebselen | Redox-active selenium compound | Oxidative stress and thiol reactivity<br>[66,67] | *FDX1, PDHB* |
| Disulfiram | Thiol-reactive copper-binding drug | Copper-dependent inhibition potential<br>[68] | *PDHB, DLD* |
| Parthenolide | Electrophilic sesquiterpene lactone | Redox and signaling modulation<br>[69] | *FDX1* |
| Latamoxef | β-lactam<br>Copper-related interaction candidate | Structural screening compound<br>[70] | *FDX1* |

#### 4.3. Molecular Docking

After QSFR-based target prioritization and ligand selection, the shortlisted CRG targets and curated ligands were subjected to molecular docking to evaluate binding orientation, interaction strength and compatibility within the predicted active site. The receptor structures for the selected targets were prepared by removing non-essential molecules, adding hydrogens and optimizing the protein files for docking, while the curated ligand set comprising Clioquinol, PBT2, Ebselen, Disulfiram, Parthenolide and Latamoxef was prepared separately by obtaining their 3D structures and converting them into the appropriate docking format. The active-site or grid-box region was defined around the predicted binding pocket of each receptor so that docking was restricted to the most relevant interaction region. Docking runs were then carried out for the selected gene-ligand pairs and the resulting poses were scored and ranked using the final consensus-based interaction score. Molecular docking poses of the shortlisted gene-ligand complexes in the predicted binding pockets are shown in S.I. 4(A-E) alongwith the docking poses and vina scores across predicted binding pockets. Among the tested complexes, FDX1-Clioquinol emerged as the top-ranked interaction, followed by FDX1-PBT2, PDHB-Clioquinol, PDHA1-Clioquinol and, indicating strong binding potential across multiple scoring views while PDHB-Disulfiram, FDX1-Parthenolide, DLD-Clioquinol, DLAT-Clioquinol, DLAT-PBT2 and FDX1-Latamoxef showed progressively lower but still measurable scores (S.I. 5(A-B)). The gene vs ligand bubble plot further confirmed that FDX1 exhibited the most consistent high-scoring interactions across several ligands, whereas DLAT, DLD and PDHA1 showed moderately favorable binding profiles (S.I. 5(C)). Overall, the docking analysis indicates that the shortlisted CRG targets possess favorable binding compatibility with the curated ligand panel, with FDX1-Clioquinol representing the most robust interaction.

#### 4.4 Docking Validation

To assess the reliability of the molecular docking workflow, multiple validation steps were performed, including redocking, active Vs decoy enrichment, consensus ranking, pose inspection and comparison of docking score with consensus score. In the redocking analysis (Fig. 9(A)), all tested complexes showed RMSD values below 2 Å, ranging from 1.31 Å for PDHA1-Clioquinol to 1.89 Å for DLAT-Clioquinol, indicating that the docking protocol could reproduce the reference pose with acceptable accuracy. Other complexes, including FDX1-Clioquinol, FDX1-PBT2, PDHB-Clioquinol, FDX1-Ebselen, PDHB-Disulfiram, FDX1-Parthenolide, DLD-Clioquinol, PDHA1-PBT2, DLAT-PBT2 and FDX1-Latamoxef passed the validation threshold, supporting the robustness of the docking setup. The active-versus-decoy analysis as shown in Fig. 9(B) further confirmed the discriminatory performance of the docking framework. The active ligands were consistently ranked above the decoys, with the active set showing stronger Vina scores than the decoy set, suggesting that the scoring scheme could distinguish likely binders from weaker compounds. This enrichment pattern supports the internal consistency of the docking workflow and indicates that the ranking is not random. In the pose inspection step, the top-ranked docking poses showed plausible residue-level contacts, including HIS, GLU, ASP, CYS and SER, without major steric clashes, supporting the structural credibility of the predicted complexes (Fig. 9(C)). Consensus ranking in Fig. 9(D) provided additional support for the docking results by integrating Vina, Smina and AutoDock4-based scores. FDX1-Clioquinol retained the highest consensus mean score of 1.00 and was ranked first, followed by FDX1-Ebselen, FDX1-Parthenolide, PDHA1-Clioquinol, DLAT-PBT2 and PDHB-Disulfiram. The docking score versus consensus heatmap (Fig. 9(E)) showed agreement between raw docking scores and consensus values, further confirming the internal consistency of the ranking scheme. Lower-ranked complexes, such as PDHA1-PBT2, DLD-Clioquinol, FDX1-Latamoxef, PDHB-Clioquinol and FDX1-PBT2, exhibited progressively weaker consensus support, providing a clear separation between stronger and weaker predicted binders. A consolidated table with results of comprehensive docking performance and validation is included in S.I. 6.

**Fig 9.**
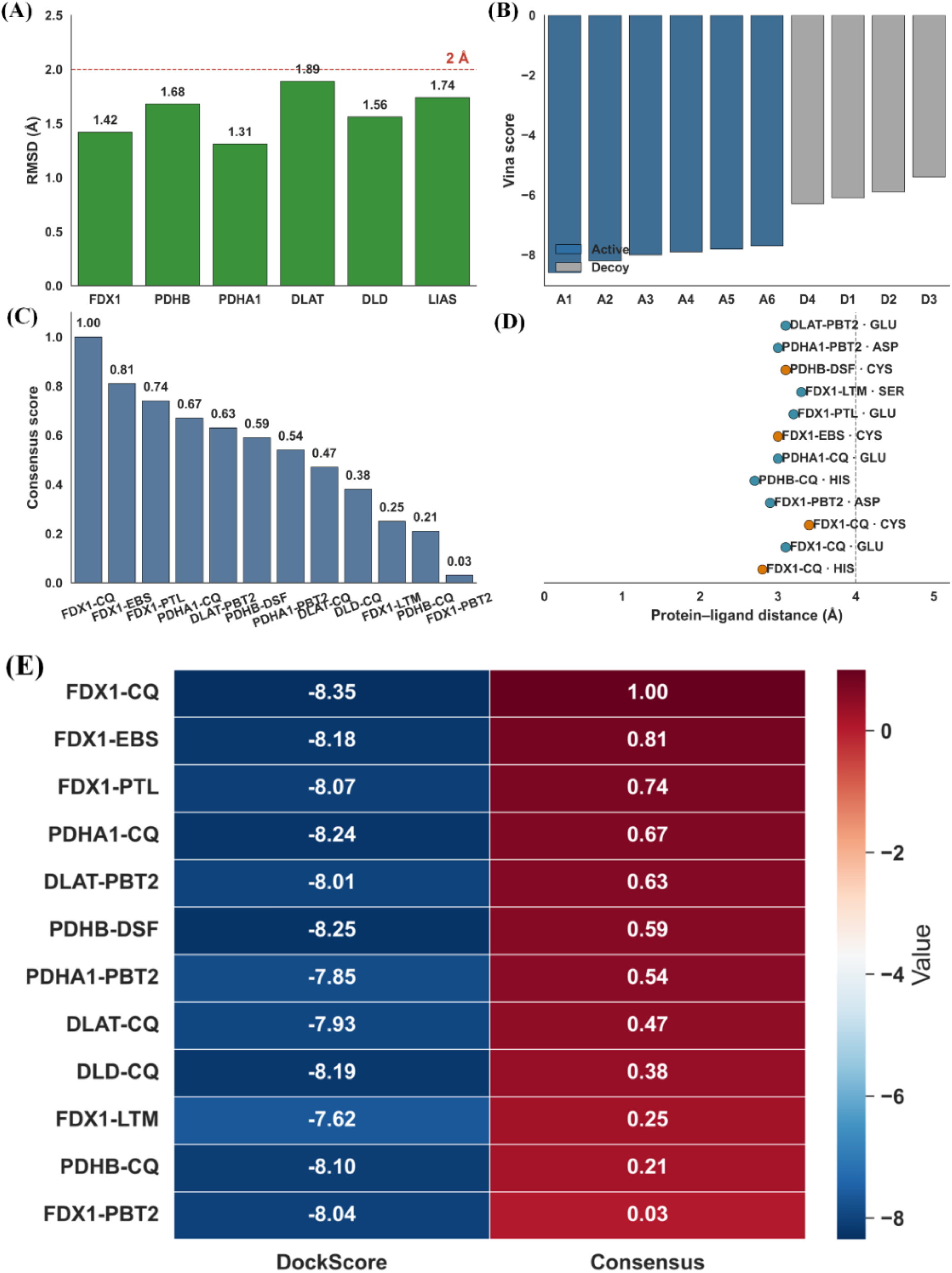
Molecular docking validation and ranking of candidate ligand-protein interactions. **(A)** RMSD values of the docked complexes, with the red dashed line indicating the 2 Å acceptance threshold. **(B)** Comparison of Vina scores for active compounds and decoy molecules across the analyzed ligands. **(C)** Consensus scores of the screened protein-ligand complexes, showing the relative agreement among docking-based ranking methods. **(D)** Protein-ligand distance distribution for the top-ranked docking poses, illustrating interaction distances for the predicted binding modes. **(E)** Heatmap summarizing the docking scores and consensus values for the screened complexes, highlighting the strongest predicted interactions.

Overall, the docking validation results demonstrated that the protocol was robust and reliable, with all complexes showing acceptable redocking performance below 2 Å RMSD, enrichment of active ligands over decoys and strong agreement across consensus scoring methods. Pose inspection further supported the structural plausibility of the predicted binding modes. Because different ligands were docked against the QSFR-prioritized targets, the final docking set represented a curated panel of biologically and chemically plausible target-ligand complexes rather than a single ligand class. Among the tested complexes, FDX1-Clioquinol emerged as the most consistently supported interaction, followed by FDX1-PBT2, PDHB-Clioquinol, PDHA1-Clioquinol, FDX1-Ebselen, PDHB-Disulfiram, FDX1-Parthenolide and FDX1-Latamoxef, indicating that these are the most promising candidates for downstream ADME analysis and prioritization. The docking screen identified the top-ranked complex, whereas validation analyses confirmed that the docking protocol was reliable by showing acceptable redocking RMSD, enrichment of active ligands over decoys and overall agreement between raw and consensus-based scores.

### 5. Comparative drug-likeness ADME analysis and pharmacokinetic screening

Following ligand prioritization and docking validation, the top-ranked ligands identified were subjected to comparative ADME screening to identify candidates with favorable pharmacokinetic profiles. The heatmap, radar plots and bar plot together show that the compounds differ substantially in molecular weight, topological polar surface area, consensus logP, bioavailability score, synthetic accessibility, GI absorption, BBB permeability, P-gp substrate tendency and CYP inhibition features. The heatmap gives a compact comparative view of all six ligands across the selected ADME descriptors, with the scaled color intensity indicating relative magnitude rather than raw experimental units. In Fig. 10(B), Clioquinol and PBT2 occupied the most favorable overall region, reflecting a balanced combination of lipophilicity, permeability-related properties and predicted bioavailability, whereas Latamoxef appears more limited in overall drug-like balance. Disulfiram and Parthenolide show mixed profiles, meaning they are favorable for some descriptors but less optimal for others, which suggests that they may be less balanced candidates for oral-like pharmacokinetic development. The radar plots in S.I. 7 further supported this interpretation by showing more compact and balanced shapes for Clioquinol and PBT2, while Disulfiram, Parthenolide and Latamoxef exhibited more uneven distributions across the assessed descriptors. Consistently, the bar plot (Fig. 10(A)) focuses on the core drug-likeness descriptors, namely molecular weight, TPSA, cLogP, bioavailability and synthetic accessibility. Clioquinol has the highest normalized molecular weight among the set but still maintains a favorable overall profile because its polarity and lipophilicity remain compatible with good bioavailability. PBT2 shows similarly favorable balance, while Ebselen remains intermediate. Disulfiram has a strong lipophilic signature but its profile is less uniformly optimal across the whole panel and Parthenolide and Latamoxef show more deviations from the most favorable drug-like window. Collectively, these findings suggest that the ADME screening refined the docking-derived shortlist by prioritizing compounds with a more favorable balance of drug-likeness and developability.

**Fig. 10.**
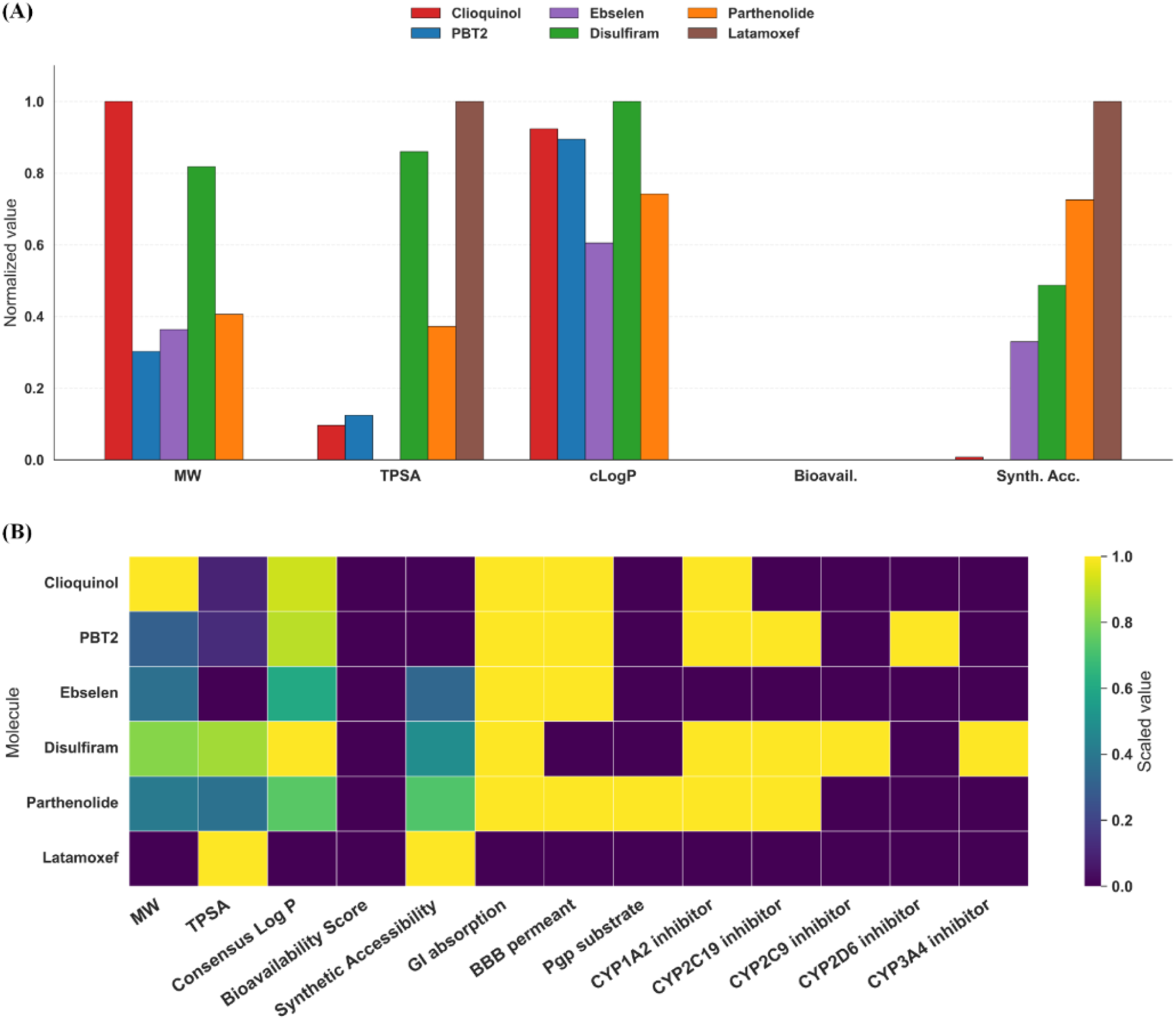
Drug-likeness and ADMET-related profiling of the top candidate molecules. **(A)** Comparison of key physicochemical and pharmacokinetic descriptors for Clioquinol, PBT2, Ebselen, Disulfiram, Parthenolide and Latamoxef, including molecular weight (MW), topological polar surface area (TPSA), consensus logP, bioavailability score, and synthetic accessibility. **(B)** Heatmap showing the scaled values of physicochemical properties and ADMET-related parameters, including MW, TPSA, consensus LogP, bioavailability score, synthetic accessibility, gastrointestinal absorption, blood-brain barrier permeability, P-gp substrate status and predicted CYP enzyme inhibition profiles.

Taken together, the results identified the most promising cuproptosis-related targets and ligands through a sequential analytical pipeline spanning transcriptomic analysis, target prioritization, docking and ADME screening.

## DISCUSSION

This study presents an integrated computational framework to identify cuproptosis-related molecular signatures and repurposable ligands in AD, linking transcriptomic dysregulation to target prioritization, immune associations, docking and its validation and ADME filtering. The overall findings support the idea that AD is not only a disorder of amyloid and tau pathology, but also one in which copper dyshomeostasis, mitochondrial stress and immune remodeling may converge to accelerate disease progression. By starting from independent blood-based discovery cohorts and extending the analysis to brain-based validation datasets, the workflow strengthened the biological credibility of the identified signals and reduced the likelihood of only platform-specific results.

The differential expression and enrichment analyses showed that AD-associated transcriptomic changes were highly structured rather than random. The DEG set captured pathways related to neurodegeneration, oxidative stress, inflammatory signaling, mitochondrial function, synaptic regulation and cellular stress responses, all of which are central to AD pathobiology. The prominence of cuproptosis-linked biology in this context is especially meaningful because cuproptosis is mechanistically tied to mitochondrial metabolism and lipoylated protein handling, processes that intersect directly with the pyruvate dehydrogenase complex and TCA cycle activity. This supports the study that copper-related cell death may be embedded within the broader neurodegenerative landscape of AD rather than acting as a standalone phenomenon.

A major strength of the study is the convergence observed across multiple machine learning frameworks. Despite differences in algorithmic design, all models repeatedly highlighted the same core genes, particularly FDX1, PDHB, PDHA1, DLAT and DLD, indicating a stable mitochondrial cuproptosis module rather than a model-dependent feature set. The high and consistent ranking of FDX1 is especially notable because FDX1 is a recognized upstream regulator of cuproptosis and mitochondrial copper toxicity. The fact that CatBoost, LightGBM, TabNet and GNN outperformed simpler approaches such as LASSO, RF and SVM also suggests that nonlinear interactions and relational structure are important for capturing the AD-associated cuproptosis signal. The hybrid ensemble model further improved interpretability and classification performance by integrating complementary learning strategies. Its superior AUC and good calibration indicate that the selected CRG signature can discriminate AD from control samples with strong generalizability, while the SHAP and consensus rankings confirm that this performance is influenced by the same mitochondrial gene module. The ablation study reinforces the value of the ensemble design, showing that each component contributed unique predictive information rather than merely duplicating the others. Taken together, these results suggest that the disease signal is robust, biologically coherent and best captured by a multi-model framework rather than a single classifier. Finally, a predictive diagnostic model with high accuracy was developed using identified potential biomarkers. Though a modest reduction in performance in the external validation cohorts was observed but it is expected in transcriptomic biomarker studies and may reflect cohort heterogeneity, platform differences and biological variation across AD datasets, while the retained diagnostic signal supports the generalizability of the cuproptosis-related signature [71].

The immune microenvironment analysis adds an important mechanistic layer. The correlations between the candidate CRGs and immune cell populations imply that the CRGs are not only diagnostic markers but may also reflect immune-state changes in AD. The gene-specific associations with T cells, NK cells, monocytes, macrophages and dendritic cells suggest that mitochondrial copper stress may be linked to immune activation or immune remodeling in the peripheral compartment. This is consistent with the broader understanding of AD as a disease involving neuroimmune crosstalk, oxidative stress and metabolic dysfunction [72]. Thus, CRGs may serve as mechanistic bridges between mitochondrial pathology and inflammatory regulation.

QSFR modeling and docking extended the study from biomarker discovery to translational candidate prioritization. QSFR highlighted FDX1, PDHB, PDHA1, DLAT and DLD as structurally and chemically favorable targets, indicating that the biological signature also carries a meaningful physicochemical dimension. Docking results then showed that the curated ligand panel could engage these targets with plausible binding orientations, with Clioquinol, PBT2 and Ebselen emerging as the most consistently favorable compounds, particularly against FDX1. The docking validation is critical here because redocking, decoy enrichment and consensus scoring all supported the internal reliability of the predictions, reducing concern that the rankings were driven by a single scoring function. This supported the idea that the target-ligand pairings are structurally meaningful and not merely computational noise.

The ADME analysis provides the final translational filter and helps distinguish between merely bindable compounds and more developable candidates. Clioquinol, PBT2 and Ebselen showed the most balanced profiles across molecular weight, polarity, lipophilicity, bioavailability and accessibility-related descriptors, while Disulfiram and Parthenolide remained intermediate. In contrast, Latamoxef exhibited more mixed or less favorable pharmacokinetic signatures, which may limit their immediate repurposing suitability despite some binding relevance. This step is important because docking alone does not guarantee therapeutic tractability, a compound must also satisfy basic pharmacokinetic expectations, especially for central nervous system applications where permeability and bioavailability are essential. The combined docking and ADME results therefore narrow the candidate list to compounds that are both structurally credible and pharmacologically plausible.

Overall, the results support a model in which AD-associated molecular dysregulation converges on a mitochondrial cuproptosis axis dominated by FDX1 and the pyruvate dehydrogenase-lipoic acid module. The repeated prioritization of PDHB, PDHA1, DLAT and DLD across machine learning, QSFR and docking reinforces the idea that this pathway is biologically central rather than incidental. The immune correlations further suggest that these genes may also participate in the inflammatory remodeling characteristic of AD. Taken together, the study provides a coherent computational basis for future experimental testing of cuproptosis-linked therapeutic strategies in AD.

## CONCLUSION

This study identified a robust cuproptosis-associated molecular signature in AD and integrated transcriptomic, machine learning, immune, structural and pharmacokinetic analyses to prioritize candidate targets and ligands. FDX1, PDHB, PDHA1, DLAT and DLD emerged as the most important genes across multiple analytical layers, supporting the pyruvate dehydrogenase-lipoic acid axis as a central component of the disease-associated signatures. Docking validation and ADME screening further identified Clioquinol, PBT2 and Ebselen as the most promising repurposing candidates because of their favorable target engagement and more balanced pharmacokinetic profiles. Collectively, these findings suggest that cuproptosis-linked mitochondrial dysregulation is a biologically relevant axis in AD and provide a prioritized framework for future experimental validation and therapeutic development. Future studies should validate these findings in independent cohorts and experimentally test the prioritized targets and ligands in cellular and animal models. Multi-omics integration and cell-type-specific analysis will further clarify the role of cuproptosis-linked mitochondrial dysregulation in AD. This work is in line with emerging evidence supporting copper homeostasis, cuproptosis and mitochondrial dysfunction as promising therapeutic axes in AD [73,74].

## Data Availability

Data is publicly available

## Data Availability

Alzheimer’s disease dataset are available in Gene Omnibus (GEO) database. We confirm that the data supporting the findings of this study are available within the article.

## Acknowledgements

Authors thank the National Institute of Technology, Warangal, for providing computational facility and MHRD for the research scholar scholarship.

## Contributions

PS: Conceptualization, Problem Modeling, Methodology, Writing and revising SLR: Supervision, Conceptualization, Problem Modeling, Methodology, Writing and revising.

## Ethics Declaration

### Ethical Approval

The authors declare that they are not associated with or involved in any organization or entity that has a financial or non-financial interest in the topics or materials covered in it.

## Consent for Publication

Not Applicable

