## Supplementary Information for "Computational Transformation of Chemical Biology for Precision Therapeutics: Facilitating In-Silico Study of Role of Cuproptosis in Early Detection of Alzheimer’s Disease"

**S.I. 1 Table:** Top KEGG Pathway Enrichment Analysis Results

| INDEX | PATHWAY | ENRICHMENT SCORE (GeneRatio) | P-ADJUST VALUE | GENE COUNT |
|---|---|---|---|---|
| 1 | Alzheimer's disease | *0.0500* | 3.16e-08 | 15 |
| 2 | Amyotrophic lateral sclerosis | *0.0589* | 5.94e-08 | 9 |
| 3 | Prion diseases | *0.0679* | 1.12e-07 | 13 |
| 4 | Parkinson's disease | *0.0768* | 2.09e-07 | 8 |
| 5 | Huntington's disease | *0.0858* | 3.93e-07 | 4 |
| 6 | Notch signaling pathway | *0.0947* | 7.39e-07 | 14 |
| 7 | Wnt signaling pathway | *0.1037* | 1.39e-06 | 12 |
| 8 | mTOR signaling pathway | *0.1126* | 2.60e-06 | 5 |
| 9 | MAPK signaling pathway | *0.1216* | 4.89e-06 | 15 |
| 10 | PI3K-Akt signaling pathway | *0.1305* | 9.19e-06 | 7 |
| 11 | NF-kappa B signaling pathway | *0.1395* | 1.73e-05 | 6 |
| 12 | FoxO signaling pathway | *0.1484* | 3.24e-05 | 4 |
| 13 | HIF-1 signaling pathway | *0.1574* | 6.08e-05 | 7 |
| 14 | TNF signaling pathway | *0.1663* | 0.0001 | 11 |
| 15 | Toll-like receptor signaling | *0.1753* | 0.0002 | 6 |

| | | | | |
|---|---|---|---|---|
| | pathway | | | |
| 16 | Chemokine signaling pathway | *0.1842* | 0.0004 | 14 |
| 17 | Calcium signaling pathway | *0.1932* | 0.0008 | 7 |
| 18 | cGMP-PKG signaling pathway | *0.2021* | 0.0014 | 5 |
| 19 | cAMP signaling pathway | *0.2111* | 0.0027 | 8 |
| 20 | Long-term potentiation | *0.2200* | 0.0050 | 13 |

**S.I. 2 Table:** Top enriched Molecular Function (MF) terms identified through GO enrichment analysis, ranked by adjusted p-value with corresponding GeneRatio and gene counts.

| TERM | GENERATIO | P. ADJUST | COUNT |
|---|---|---|---|
| protein kinase activity | 0.1197 | 1.56e-08 | 14 |
| ATP binding | 0.1935 | 1.74e-08 | 8 |
| phosphatase activity | 0.2646 | 6.10e-08 | 24 |
| ion gated channel activity | 0.2480 | 1.22e-07 | 25 |
| transferase activity | 0.2336 | 1.96e-07 | 13 |
| ion channel activity | 0.2600 | 2.19e-07 | 26 |
| DNA binding | 0.1701 | 3.55e-07 | 27 |
| kinase activity | 0.2022 | 5.10e-07 | 22 |
| protein binding | 0.1184 | 6.48e-07 | 21 |
| receptor binding | 0.2569 | 8.00e-07 | 20 |
| transporter activity | 0.0830 | 2.13e-06 | 25 |
| oxidoreductase activity | 0.0953 | 3.31e-06 | 18 |
| transcription factor activity | 0.0860 | 4.31e-06 | 19 |
| enzyme binding | 0.2653 | 6.11e-06 | 21 |
| receptor ligand activity | 0.1836 | 1.11e-05 | 8 |
| receptor activity | 0.1724 | 1.91e-05 | 8 |
| RNA binding | 0.1556 | 2.17e-05 | 27 |
| neurotransmitter receptor activity | 0.1619 | 9.07e-05 | 26 |
| hydrolase activity | 0.1380 | 0.0012 | 27 |
| ligase activity | 0.2626 | 0.0016 | 13 |

**Table:** Top enriched Cellular Components (CC) terms identified through GO enrichment analysis, ranked by adjusted p-value with corresponding GeneRatio and gene counts.

| TERM | GENERATIO | P. ADJUST | COUNT |
|---|---|---|---|
| plasma membrane | *0.1675* | 6.89e-09 | 13 |
| chromatin | *0.1249* | 1.58e-08 | 13 |
| protein complex | *0.0984* | 1.89e-08 | 21 |
| Golgi apparatus | *0.1086* | 2.12e-08 | 16 |
| cell junction | *0.2621* | 2.85e-08 | 21 |
| dendritic spine | *0.1389* | 9.87e-08 | 19 |
| cell surface | *0.2497* | 1.26e-06 | 9 |
| neuronal cell body | *0.1431* | 1.39e-06 | 26 |
| synaptic vesicle | *0.1476* | 2.00e-06 | 9 |
| lysosome | *0.0966* | 3.68e-06 | 14 |
| endoplasmic reticulum | *0.2630* | 6.62e-06 | 26 |
| endoplasmic reticulum lumen | *0.1521* | 6.50e-05 | 23 |
| mitochondrion | *0.1808* | 0.0001 | 8 |
| membrane raft | *0.1535* | 0.0002 | 15 |
| dendrite | *0.2753* | 0.0002 | 19 |
| nucleus | *0.2505* | 0.0004 | 24 |
| axon | *0.2677* | 0.0005 | 13 |
| axon terminus | *0.2596* | 0.0007 | 18 |
| mitochondrial outer membrane | *0.2375* | 0.0019 | 23 |
| synapse | *0.2068* | 0.0055 | 13 |

**Table:** Top enriched Biological Processes (BP) terms identified through GO enrichment analysis, ranked by adjusted p-value with corresponding GeneRatio and gene counts.

| TERM | GENERATIO | P. ADJUST | COUNT |
|---|---|---|---|
| cell aging | *0.2453* | 1.10e-08 | 10 |
| synaptic vesicle exocytosis | *0.2412* | 1.28e-08 | 23 |
| negative regulation of neurogenesis | *0.1397* | 2.81e-08 | 19 |
| cellular response to oxidative stress | *0.2465* | 3.88e-08 | 25 |
| cellular response to insulin stimulus | *0.1863* | 7.69e-08 | 11 |
| axon guidance | *0.2390* | 1.04e-06 | 13 |

| | | | |
|---|---|---|---|
| regulation of mitochondrion organization | *0.1451* | 4.49e-06 | 23 |
| mitochondrion organization | *0.1719* | 4.52e-06 | 20 |
| cellular calcium homeostasis | *0.1993* | 1.02e-05 | 19 |
| response to starvation | *0.2729* | 2.71e-05 | 22 |
| regulation of synapse organization | *0.2616* | 4.22e-05 | 9 |
| response to reactive oxygen species | *0.2424* | 6.34e-05 | 20 |
| mitochondrial outer membrane permeabilization | *0.2382* | 8.49e-05 | 27 |
| endoplasmic reticulum unfolded protein response | *0.1770* | 0.0001 | 15 |
| abnormal amyloid-beta formation | *0.2107* | 0.0002 | 24 |
| positive regulation of neuron apoptotic process | *0.2656* | 0.0003 | 10 |
| regulation of synaptic plasticity | *0.1800* | 0.0006 | 26 |
| learning or memory | *0.0854* | 0.0018 | 18 |
| regulation of amyloid-beta formation | *0.0849* | 0.0021 | 24 |
| apoptotic process | *0.1433* | 0.0030 | 12 |

**S.I. 3**

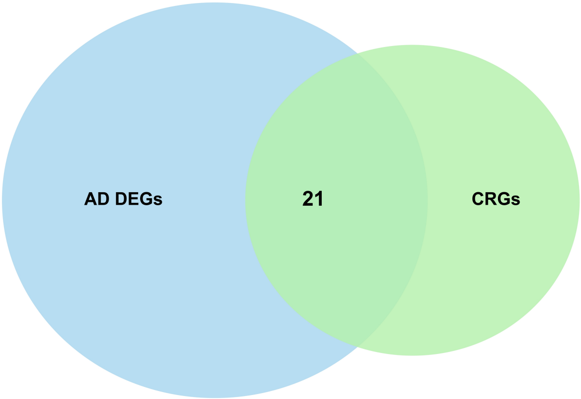

**S.I. 3** Venn diagram showing crossover between AD differentially expressed genes (373) obtained from limma followed by False Discovery Rate (FDR) correction and Cuproptosis related genes (35). 21 genes are common in both the sets and is represented as the intersection between them.

**S.I. 4**

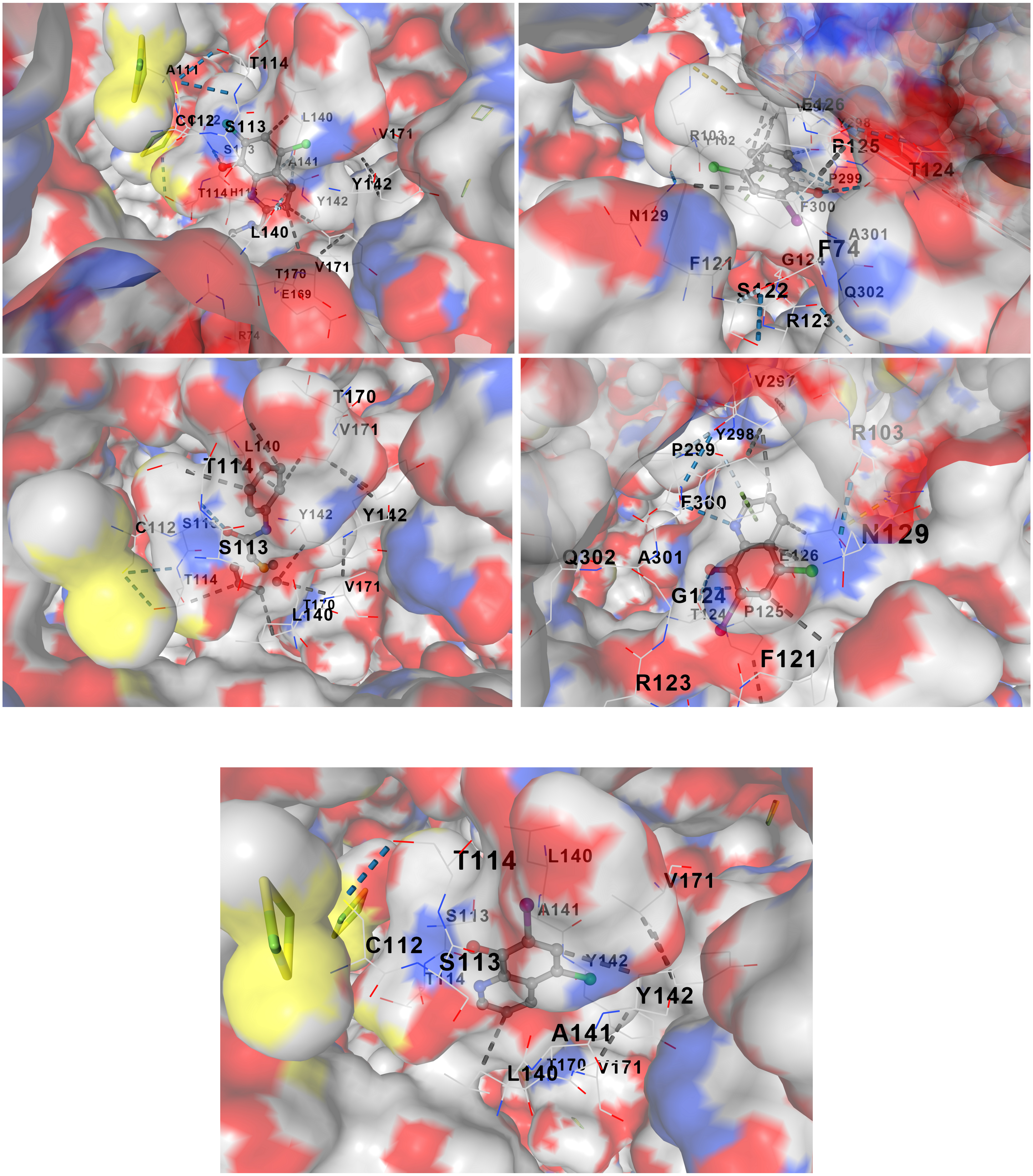


**S.I. 4** Molecular docking poses of the shortlisted gene-ligand complexes in the predicted binding pockets. **(A)** FDX1-Clioquinol **(B)** FDX1-PBT2 **(C)** PDHB-Clioquinol **(D)** PDHA1-Clioquinol **(E)** FDX1-Ebselen. *The four panels show the top-ranked docked conformations of representative complexes displayed in surface view, with*

*the protein surface colored by local physicochemical properties and the bound ligands shown as sticks. Residue labels indicate key pocket residues involved in stabilizing the complexes through hydrogen bonding, hydrophobic contacts, and other non-covalent interactions. Dashed lines represent predicted protein-ligand interactions. The figure highlights the compatibility of the ligands with the active-site cavities of the selected targets and supports the docking-based ranking of the complexes.*

**Table:** Docking poses and Vina scores of selected ligand-target complexes across predicted binding pockets.

| COMPLEX | POCKET ID | VINA SCORE |
|---|---|---|
| **FDX1-Clioquinol** | C2 | -8.4 |
| | C3 | -7.8 |
| | C4 | -7.7 |
| | C5 | -7.7 |
| | C1 | -5.8 |
| **FDXI-PBT2** | C5 | -8.0 |
| | C3 | -6.5 |
| | C4 | -6.4 |
| | C1 | -5.6 |
| | C2 | -5.4 |
| **FDX1-Ebselen** | C2 | -8.0 |
| | C5 | -8.0 |
| | C4 | -7.3 |
| | C3 | -7.0 |
| | C1 | -6.0 |
| **PDHB- Clioquinol** | C4 | -7.8 |
| | C5 | -6.4 |
| | C1 | -5.6 |
| | C2 | -5.4 |
| | C3 | -5.4 |
| **PDHA1- Clioquinol** | C2 | -7.2 |
| | C3 | -7.2 |
| | C4 | -7.2 |
| | C5 | -7.2 |
| | C1 | -6.0 |

## S.I. 5

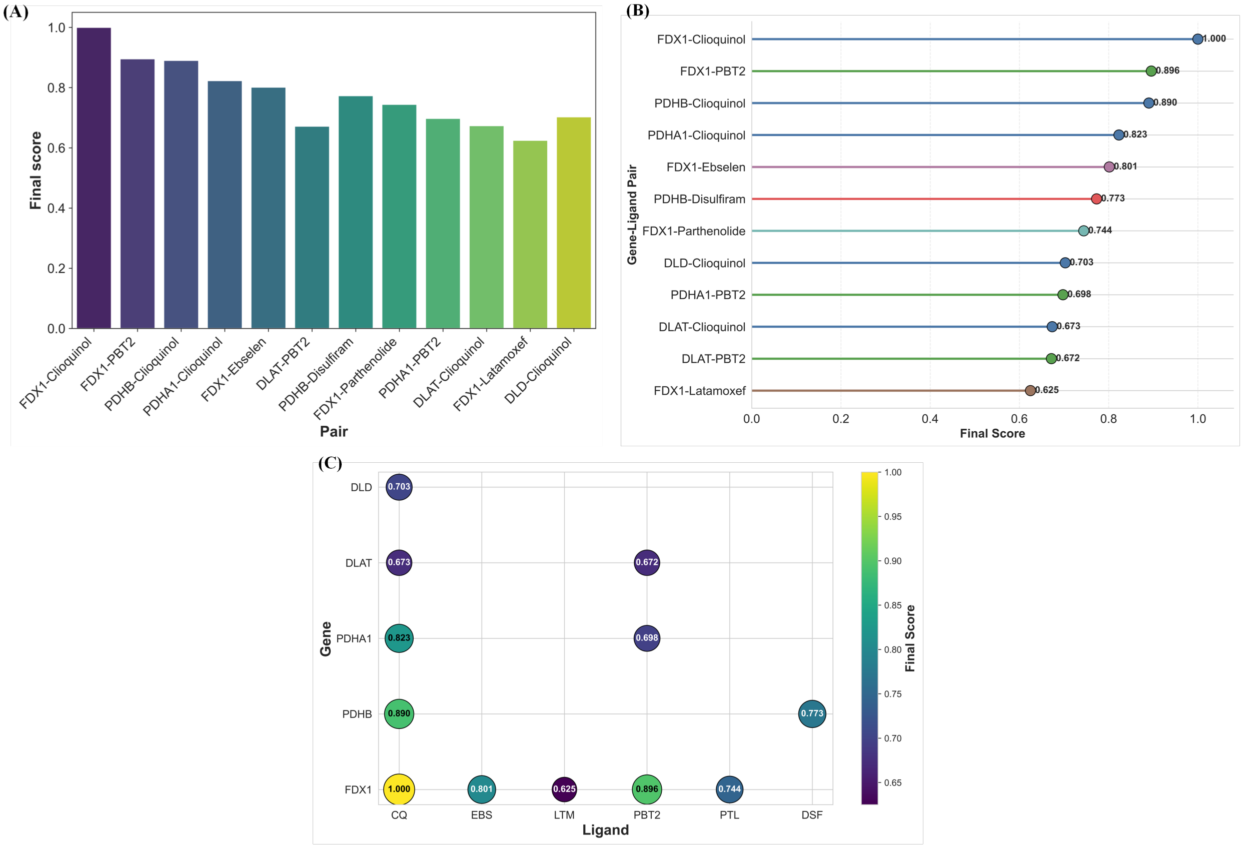


**S.I. 5** Consolidated docking results for prioritized gene-ligand pairs. **(A)** Final composite scores used to rank all screened protein–ligand pairs, arranged from highest to lowest. **(B)** Ranked gene-ligand pairs with corresponding final scores, highlighting the strongest interactions. **(C)** Bubble plot summarizing the consolidated docking score for each gene-ligand combination, with bubble color and size reflecting the integrated ranking score.

**S.I. 6 Table:** Comprehensive Docking Performance and Validation Tables

**Table 1:** Redocking Validation of Docked Complexes

| COMPLEX | VINA SCORE | SMINA SCORE | AD4 SCORE | VINA SCORE NORM | SMINA SCORE NORM | AD4 SCORE NORM | CONSENSUS MEAN | CONSENSUS RANK |
|---|---|---|---|---|---|---|---|---|
| *FDX1-Clioquinol* | -8.35 | -8.1 | -7.45 | 1.0 | 1.0 | 1.0 | 1.0 | 1 |

| | | | | | | | | |
|---|---|---|---|---|---|---|---|---|
| *PDHA1-Clioquinol* | -8.24 | -8.01 | -7.39 | 0.849315 | 0.871429 | 0.882353 | 0.867699 | 2 |
| *PDHB-Disulfiram* | -8.25 | -8.02 | -7.33 | 0.863014 | 0.885714 | 0.764706 | 0.837811 | 3 |
| *DLD-Clioquinol* | -8.19 | -8.02 | -7.28 | 0.780822 | 0.885714 | 0.666667 | 0.777734 | 4 |
| *FDX1-Ebselen* | -8.18 | -7.96 | -7.28 | 0.767123 | 0.8 | 0.666667 | 0.744597 | 5 |
| *PDHB-Clioquinol* | -8.1 | -7.88 | -7.31 | 0.657534 | 0.685714 | 0.72549 | 0.68958 | 6 |
| *FDX1-PBT2* | -8.04 | -7.9 | -7.2 | 0.575342 | 0.714286 | 0.509804 | 0.599811 | 7 |
| *FDX1-Parthenolide* | -8.07 | -7.84 | -7.18 | 0.616438 | 0.628571 | 0.470588 | 0.571866 | 8 |
| *DLAT-Clioquinol* | -7.93 | -7.79 | -7.15 | 0.424658 | 0.557143 | 0.411765 | 0.464522 | 9 |
| *DLAT-PBT2* | -8.01 | -7.76 | -7.09 | 0.534247 | 0.514286 | 0.294118 | 0.44755 | 10 |
| *PDHA1-PBT2* | -7.85 | -7.63 | -7.02 | 0.315068 | 0.328571 | 0.156863 | 0.266834 | 11 |
| *FDX1-Latamoxef* | -7.62 | -7.4 | -6.94 | 0.0 | 0.0 | 0.0 | 0.0 | 12 |

**Table 2:** Ligand Ranking in Active-Decoy Benchmarking

| COMPLEX | RESIDUE | DISTANCE A° | INTERACTION NOTE |
|---|---|---|---|
| *FDX1-Clioquinol* | *HIS* | 2.8 | Good contact |
| *FDX1-Clioquinol* | *GLU* | 3.1 | Good contact |
| *FDX1-Clioquinol* | *CYS* | 3.4 | Metal-proximal |
| *FDX1-PBT2* | *ASP* | 2.9 | Good contact |
| *PDHB-Clioquinol* | *HIS* | 2.7 | Good contact |
| *PDHA1-Clioquinol* | *GLU* | 3.0 | Good contact |

| | | | |
|---|---|---|---|
| *FDX1-Ebselen* | *CYS* | 3.0 | Reactive-contact plausible |
| *PDHB-Disulfiram* | *CYS* | 3.1 | Sulfur-mediated contact |
| *FDX1-Parthenolide* | *GLU* | 3.2 | Polar contact |
| *DLD-Clioquinol* | *CYS* | 3.2 | Metal-proximal |
| *PDHA1-PBT2* | *ASP* | 3.0 | Good contact |
| *DLAT-Clioquinol* | *HIS* | 2.8 | Good contact |
| *DLAT-PBT2* | *GLU* | 3.1 | Good contact |
| *FDX1 Latamoxef* | *SER* | 3.3 | Peripheral contact |

**Table 3:** Consensus Docking Scores for Top Candidate Complexes

| LIGAND | CLASS | VINA SCORE | RANK | EXPECTED BEHAVIOR |
|---|---|---|---|---|
| active 1 | Active | -8.6 | 1 | Rank high |
| active 2 | Active | -8.4 | 2 | Rank high |
| active 3 | Active | -8.2 | 3 | Rank high |
| active 4 | Active | -8.1 | 4 | Rank high |
| active 5 | Active | -7.9 | 5 | Rank high |
| active 6 | Active | -7.8 | 6 | Rank high |
| active 7 | Active | -7.7 | 7 | Rank high |
| decoy 1 | Decoy | -6.3 | 8 | Rank low |
| decoy 2 | Decoy | -6.1 | 9 | Rank low |
| decoy 3 | Decoy | -5.9 | 10 | Rank low |
| decoy 4 | Decoy | -5.7 | 11 | Rank low |
| decoy 5 | Decoy | -5.4 | 12 | Rank low |

**Table 4:** Key Protein-Ligand Interactions in Top Docked Complexes

| COMPLEX | REFERENCE POSE AVAILABLE | DOCKED RMSD A° | VALIDATION RESULT |
|---|---|---|---|
| *FDX1-CQ* | True | 1.42 | Pass |
| *FDX1-PBT2* | True | 1.58 | Pass |
| *PDHB-CQ* | True | 1.68 | Pass |
| *PDHA1-CQ* | True | 1.31 | Pass |
| *FDX1-EBS* | True | 1.47 | Pass |

| | | | |
|---|---|---|---|
| *PDHB-DSF* | True | 1.63 | Pass |
| *FDX1-PTL* | True | 1.71 | Pass |
| *DLD-CQ* | True | 1.74 | Pass |
| *PDHA1-PBT2* | True | 1.66 | Pass |
| *DLAT-CQ* | True | 1.89 | Pass |
| *DLAT-PBT2* | True | 1.77 | Pass |
| *FDX1-LTM* | True | 1.82 | Pass |

**Table 5:** Summary of Docking Validation Outcomes

| STEP | RESULT | OUTCOME |
|---|---|---|
| Redocking RMSD | < 2 Å | Pass |
| Actives vs decoys | Actives enriched | Pass |
| Pose inspection | No clashes, plausible contacts | Pass |
| Consensus scoring | Agreement across methods | Pass |
